# Understanding how the design and implementation of Online Consultations influence primary care outcomes: Systematic review of evidence with recommendations for designers, providers, and researchers

**DOI:** 10.1101/2022.02.21.22271185

**Authors:** Sarah Darley, Tessa Coulson, Niels Peek, Susan Moschogianis, Sabine N van der Veer, David C Wong, Benjamin C Brown

## Abstract

**Background:** Online consultations (OCs) allow patients to contact their care provider online, and have been promoted as a way to address increasing workload and decreasing workforce capacity in primary care. Globally, OCs have been rolled out rapidly due to policy initiatives and the COVID-19 pandemic, though there is a lack of evidence regarding how their design and implementation influence care outcomes.

**Objective:** Informed by existing theories, synthesise quantitative and qualitative research on: 1) outcomes of OCs in primary care; 2) how these are influenced by OC system design and implementation.

**Methods:** We searched Ovid Medline, Embase, Web of Science, Scopus, NTIS, HMIC, and ZETOC from 2010 to November 2021. We included quantitative and qualitative studies of real-world OC use in primary care, written in English, and published 2010 onwards. Quantitative data were transformed into qualitative themes. For objective 1 we used thematic synthesis informed by the Institute of Medicine’s domains of healthcare quality. For objective 2 we used Framework Analysis informed by the NASSS framework and Realistic Evaluation. Critical appraisal was conducted using the Mixed Methods Appraisal Tool and strength of evidence judged using GRADE-CERQual.

**Results:** We synthesised 62 studies (quantitative n=32, qualitative n=12, mixed methods n=18) in nine countries covering 30 unique OC systems, 13 of which used Artificial Intelligence (AI). Twenty-six were published in 2020 onwards, and 11 were post-COVID-19. There was no quantitative evidence for negative impacts of OCs on patient safety, and qualitative studies suggested perceptions of OC safety varied. Some participants believed OCs improved safety, particularly when patients could describe their queries using unstructured free-text. Staff workload decreased when sufficient resources were allocated to implement OCs, and patients used them for simple problems or could describe their queries using free-text. Staff workload increased when OCs were not integrated with other software or organisational workflows, and patients used them for complex queries. OC systems that required patients to describe their queries using multiple choice questionnaires (MCQs) increased workload for both them and staff. Health costs were reduced when patients used OCs for simple queries, and increased when used for complex ones. Patients using OCs were more likely to be female, younger, native speakers, with higher socioeconomic status than those not using OCs. However, OCs increased primary care access for patients with mental health conditions, verbal communication difficulties, and barriers to attending in-person appointments. Access also increased by providing a timely response to patients’ queries. Patient satisfaction increased when using OCs due to better primary care access, though could decrease when using MCQ formats.

**Conclusions:** This is the first theoretically-informed synthesis of research on OCs in primary care, and includes studies conducted during COVID-19. It contributes new knowledge that in addition to producing positive outcomes such as increased access and patient satisfaction, they can also have negative outcomes such as increased workload and costs. These negative outcomes can be mitigated by appropriate OC system design (e.g. free-text format), incorporating advanced technologies (e.g. AI), and integration into technical and organisational workflows (e.g. timely responses).

**Study protocol:** PROSPERO (CRD42020191802).

## Introduction

Online consultation (OC) systems allow patients to contact their healthcare provider over the internet to ask health-related questions and report symptoms [1]. Their query may then be resolved either by a written response, telephone call, video consultation, or in-person visit. Communication between provider and patient through OC systems may therefore be synchronous (real-time e.g. via video) or asynchronous (questions and responses sent at different times e.g. via written message). Many terms are used to describe this type of technology including ‘e-consultation’, ‘e-visit’, and ‘online triage’ (Appendix 1) – in this review we refer to them all as ‘OCs’. We distinguish OCs from ‘symptom checkers’ [2] and other self-service systems that typically do not directly facilitate communication with a human healthcare provider, and from patient portals [3], which may include generic email or secure messaging functionality.

OCs are considered by policymakers in many countries as a way to address the increasing workload and decreasing workforce capacity in primary care [4-9], whilst still meeting patient expectations and improving access [10]. However, they have the potential to exacerbate health inequities [11, 12], and increase inappropriate antibiotic prescriptions [13]. Furthermore, there are widely recognised challenges in initiating and sustaining adoption of new technologies in primary care [14].

Although symptom checkers (e.g. [2, 15]) and patient portals (e.g. [3, 16, 17]) have been well studied, only a small number of evidence syntheses directly relevant to OCs have been published: one systematic review of 57 articles on delivering ‘e-consultation’ in primary care largely focused on generic stand-alone applications such as email and video (n=39/57, 68%) [18]; a scoping review of ‘online triage tools’ included 13 papers, four of which (31%) were non-empirical (e.g. opinion pieces) [19]; and a review of 17 studies of ‘intelligent online triage tools’ focused only on those that use ‘artificial intelligence’ (AI) [20].

Since these syntheses were conducted, OCs have gained wider traction in clinical practice around the world: they have been indispensable in helping manage patients remotely to minimise the spread of COVID-19 [21, 22], and English primary care providers have been mandated to offer OCs for all patients since April 2020 [23]. Moreover, OC system product design has progressed significantly to become more specialised and technologically advanced [24], with several more empirical research studies published on their use (e.g. [25-47]).

Given this rapid scale-up and increase in diversity and complexity of OCs, further insight is needed into their outcomes on healthcare. Previous reviews have not reported design or implementation details of the OCs they studied (e.g. [18-20]), despite their importance in understanding the causal mechanisms of how they impact outcomes [48]. In this paper, we systematically review and synthesise the empirical quantitative and qualitative literature in a theoretically-informed way to address this knowledge gap.

## Objectives

Informed by existing theories, synthesise quantitative and qualitative research on:

1. Outcomes of OCs in primary care.
2. How these are influenced by OC system design and implementation.

## Methods

### Study design

We consider OCs as complex interventions and therefore synthesised both quantitative and qualitative evidence to understand their impacts in specific contexts [49]. We did not perform a meta-analysis due to the heterogeneous and non-randomised nature of included studies [50]. We followed the Preferred Reporting Items for Systematic Reviews and Meta-Analyses (PRISMA) statement [51].

### Inclusion criteria

Papers that met the following criteria were included: empirical studies employing quantitative and/or qualitative methods to examine the real-world use of OCs in primary care in any country, written in English, and published in 2010 or later. We excluded news articles, opinion pieces, literature reviews, non-English language articles, and literature published before 2010.

We defined OCs as digital interventions that allow patients to contact their primary care provider by inputting ‘queries’ into healthcare-specific online forms [1]. We included symptom checkers and similar self-service systems (e.g. [29]) if at least one of their outcomes directly facilitated contact with a primary care health professional. We included patient portals if they had secure messaging functionality that used healthcare-specific forms (e.g. [29]). We excluded stand-alone generic communication technologies, such as email or video conferencing software.

### Search strategy

We searched databases Ovid Medline, Embase, Web of Science, and Scopus during July 2020 (Appendix 2). Our search strategy was developed from our scoping searches of the literature, and drew on search strategies used in related literature reviews [18, 19]. We searched NTIS (National Technical Information Service), HMIC (Health Management Information Consortium), and ZETOC to find relevant grey literature, conference proceedings, and theses. We found further literature through citation mapping and reference lists of relevant papers, searching during August and September 2020. SD and TC independently screened titles and abstracts, then full papers for eligibility, resolving differences through discussion at each stage. All literature searches were re-run by SD in November 2021.

### Data extraction and quality appraisal

We extracted data from included papers as verbatim text, capturing study characteristics (e.g. research design, study setting) and key findings relevant to our research objectives based on the NASSS framework [52] (Appendix 3). We used the NASSS in order to capture ‘a rich, contextualised narrative of technology-supported change efforts and the numerous interacting influences that help explain its successes, failures, and unexpected events’ [53]. Methodological quality of studies was assessed using the Mixed Methods Appraisal Tool (MMAT), which is designed for qualitative, quantitative, and mixed methods studies [54]. We scored each paper using recommended quintile percentages as cut-offs, and considered any paper scoring at least 60% as ‘good’ quality [55]. SD and TC extracted data for ten papers independently, which confirmed high inter-rater agreement. Following this, SD extracted data from the remaining papers, which were checked by TC.

### Data synthesis

Data were imported into Nvivo (v12) [56] for synthesis. In order to integrate both quantitative and qualitative data, during data synthesis quantitative data were transformed into qualitative themes (‘qualitising’) [57].

For objective 1, we considered ‘outcomes’ as consequences of using OCs that could relate to patients, primary care staff, or the wider system [48]. We used thematic synthesis [58], which involved SD and TC coding the text from the data extraction forms independently line-by-line, developing higher level themes through regular discussion [58]. Outcomes were synthesised inductively, with emerging themes mapped to the six Institute of Medicine (IOM) domains of healthcare quality [59]: Safe (avoiding harm to patients from care that is intended to help), Effective (providing care based on scientific knowledge to produce better clinical outcomes), Patient-centred (care that is respectful and responsive), Timely (reducing waits and delays for those who receive and give care), Efficient (avoiding waste), and Equitable (care that does not vary in quality because of personal characteristics) [59]. Our emergent findings suggested OCs had both positive and negative outcomes, and therefore theme descriptions were edited to be neutral (e.g. Safe→Safety, Efficient→Efficiency). Findings for the Effective domain were rated as low confidence and are therefore not presented in our main results.

For objective 2, we considered OC ‘design’ as material properties of an OC such as features and functionality [52], and ‘implementation’ as the way an OC had been introduced and used in a particular context [48]. As a design feature, we considered AI as the ability of machines to ‘mimic human intelligence as characterised by behaviours such as cognitive ability, memory, learning, and decision making’ [60]. We synthesised extracted data using Framework Analysis [61], which involved SD and TC reading and re-reading each data extraction form, then coding them line-by-line independently: both deductively using domains from the NASSS framework [52] for high-level themes, and inductively by identifying additional subthemes. Through discussion, SD and TC summarised the findings into five high-level themes: Condition (health condition, illness OC is used for), Technology (material properties of OC, required knowledge for use), Adopters (staff, patients, carers expected to use OC), Organisation (extent of work needed for implementation of OC, capacity, and readiness), and Wider System (policy context) [52]. Two NASSS domains, Value Proposition (value of OC to the developer, patients, and healthcare system) and Embedding and Adaptation over Time (learning and adaptation to changing contexts), had limited applicability to our findings and were not included in the final synthesis. Informed by Realistic Evaluation [48], we considered our themes as contextual factors, and identified patterns of explanations for how each led to the outcomes from objective 1 (i.e. ‘causal mechanisms’). We used visual mapping to identify commonalities and discordances in causal mechanisms – firstly within individual papers, then secondly across papers [61]. Where there were discordances, we explored potential explanations where possible (e.g. related to study setting).

Strength and quality of our findings for both objective 1 and 2 was assessed using the GRADE-CERQual method [62]. This accounts for: methodological limitations of contributing papers (according to MMAT assessments); relevance to the review question; coherence of the finding; and adequacy of its supporting data [62]. Confidence in each finding was designated as either high, moderate, low, or very low. At each stage of analysis, findings were discussed and agreed with the wider study team. Author BCB reviewed all coded verbatim excerpts from papers included in the final synthesis.

## Results

### Descriptive summary

We synthesised 62 papers (Figure 1), including 51 journal papers (e.g. [26]), seven evaluation reports (e.g.[63]), three conference papers (e.g. [64]), and one Master’s degree thesis [65]. Studies were quantitative (n=32, 52%), qualitative (n=12, 19%), and mixed methods (n=18, 29%), and analysed data from patients (16 qualitative, 17 quantitative studies), staff (22 qualitative studies, 9 quantitative studies), and clinical systems (32 quantitative studies). All were set in one of nine high-income countries, with the majority from the US (n=20) and UK (n=20; Appendix 4). Twenty-six (42%) studies were published in 2020 or later, and 11 studies (18%) were conducted after the start of the COVID-19 pandemic. Examples of excluded studies are those that focus on stand-alone video consultations (e.g. [66]), involved communication between physicians and not patients (e.g. [67]), and those not based in primary care (e.g. [68]).

Fifty two studies reported levels of OC adoption by patients and staff, out of which 32 (62%; 52% of all studies) were described as ‘low’ by study authors (e.g. [69]). OCs were adopted at a high rate in 33 (63%; 53% of all) studies (e.g. [70]), including high rates of adoption by certain patient groups even when overall OC adoption in the study was low (e.g. [71]).

Included papers described 30 OC systems summarised in Table 1 and detailed in Appendix 5. In 16 papers, the OC system was described sufficiently to meet our inclusion criteria but not enough detail to determine specific design features. Of the 30 OCs described, the majority (n=23, 77%) offered asynchronous two-way written communication between patients and staff i.e. text-based messages sent at different times (e.g. [72]), with a few (n=4, 13%) also offering synchronous real-time communication by video (e.g. [25]). No OCs provided synchronous text-based communication. Four (13%) did not provide functionality for staff to reply to patients via the system (i.e. one-way communication only e.g. [71]). Eleven (37%) required patients to describe their queries solely via multiple choice questionnaires (MCQs; e.g. [73]) compared to four (13%) that solely required patients to describe their queries using unstructured free-text (e.g. [31]). Twelve (40%) had a hybrid approach of primarily using MCQs with the option for patients to enter additional free-text (e.g.[74]). No free-text OCs offered optional MCQs. Eight (27%) OC systems integrated with the electronic health record (EHR; e.g.[33]) and one (3%) allowed patients to schedule telephone or in-person appointments with health care professionals themselves (e.g. [29]).

**Table 1:** OC Features

| OC system features |  | Count (%)* |
| --- | --- | --- |
| **Communication mode** | Asynchronous two-way communication | 23 (77%) |
|  | Synchronous real-time communication | 4 (13%) |
|  | One-way communication only | 4 (13%) |
|  | Unclear | 3 (10%) |
| **Patient query format** | Multiple choice questionnaires only | 11 (37%) |
|  | Unstructured free-text only | 4 (13%) |
|  | Multiple choice questionnaires with optional free-text | 12 (40%) |
|  | Unclear | 3 (10%) |
| **Integration with other software** | Electronic health record | 8 (27%) |
|  | Appointment scheduling | 1 (3%) |
|  | No integration | 22 (73%) |
| **Artificial intelligence function** | Adapting patient questions during query submission | 10 (33%) |
|  | Prioritising patient queries based on clinical urgency | 4 (13%) |
|  | Signposting patients to the most appropriate care provider | 3 (10%) |
|  | No artificial intelligence | 17 (57%) |
| **Artificial intelligence method** | Pre-programmed logic and algorithms | 10 (33%) |
|  | Unclear | 3 (10%) |

\*Count of OC systems described in detail (n=30). Categories may add up to more than 30 because OC systems may have more than one feature in a category.

Thirteen MCQ-based OC systems exhibited three types of AI: 1) adapting the questions they asked patients as they submitted their query in response to their answers so far (n=10, 33%; e.g. [75]); 2) prioritising patient queries based on clinical urgency (n=4, 13%; e.g. [29]); and 3) signposting patients to an appropriate care provider based on their query, such as self-care, primary care, or emergency department (n=3, 10%; e.g. [43]). These were mostly powered by pre-programmed logic and “algorithms” (n=10, 33%; e.g. [29]), with the exact AI methodology unclear in the remainder (n=3, 10%; e.g. [76]).

The methodological quality of most papers (n=41, 66%) was ‘good’ (i.e. 60% or above according to MMAT [77]; Appendix 6). Common limitations included lack of detail on whether the OC was administered as intended (e.g. [78]), and small sample sizes (e.g. [28]).

### Synthesis

To maintain readability we present only moderate and high confidence findings, and provide only one example reference per finding. Tables 2 and 3 provide all references, and specify whether findings are qualitative or quantitative. Appendices 7 and 8 detail low confidence findings. Appendices 9 and 10 provide exemplar data.

**Table 2.** Outcomes of Online Consultations in primary care

| Theme | Subtheme |
| --- | --- |
| **Safety** (harm to patients) | **Decreased patient safety (qualitative)**<br>**Description:** Patient and staff perceptions that OCs worsened patient safety.<br>**CERQual rating:** High<br>**References:** [27, 28, 30, 36, 38, 41, 42, 45, 63, 65, 74, 82, 83, 89-92], n=17 |
|  | **Neutral-increased patient safety (qualitative and quantitative)**<br>**Description:** No quantitative evidence of negative impacts on patient safety, with clinician and patient perceptions that OCs improved patient safety.<br>**CERQual rating:** High<br>**References:** [28-30, 32-34, 37, 38, 40, 41, 44, 46, 65, 71-73, 78-81, 83, 87, 93-95], n=25 |
| **Timeliness** (reducing waits and delays) | **Increased access (qualitative and quantitative)**<br>**Description:** Easier and more convenient for patients to contact their primary care provider, and quicker to communicate with a health professional.<br>**CERQual rating:** High<br>**References:** [27, 28, 30-33, 37, 39-42, 44, 47, 63, 65, 71, 74, 76, 78, 81-84, 86, 87, 89-91, 94], n=29 |
| **Efficiency**<br>(avoiding waste) | **Decreased workload (qualitative and quantitative)**<br>**Description:** Less work for staff and patients to provide and receive care, respectively.<br>**CERQual rating:** High<br>**References:** [28-33, 35-38, 41, 44, 46, 47, 63, 65, 71, 73, 74, 76, 78-84, 86, 87, 89, 92-94], n=33 |
|  | **Increased workload (qualitative and quantitative)**<br>**Description:** More work for staff and patients to provide and receive care, respectively.<br>**CERQual rating:** High<br>**References:** [25, 28, 30, 31, 33, 37, 38, 43-45, 47, 63, 65, 69-71, 76, 78, 80-84, 86-89, 91, 96], n=29 |
|  | **Decreased costs (qualitative and quantitative)**<br>**Description:** Lower costs for the healthcare system and patients to provide and receive care, respectively.<br>**CERQual rating:** High<br>**References:** [31, 32, 35, 36, 38, 41, 63, 73, 76, 78, 79, 81-83, 87, 89, 94, 95, 97, 98], n=20 |
|  | **Increased costs (qualitative and quantitative)**<br>**Description:** Higher costs for the healthcare system.<br>**CERQual rating:** High<br>**References:** [38, 41, 70, 81-84, 88, 89], n=9 |
| **Equitable**<br>(variation because of personal characteristics) | **Decreased equity (qualitative and quantitative)**<br>**Description:** OC use variation based on patient characteristics.<br>**CERQual rating:** High<br>**References:** [25, 32, 34, 35, 41-43, 47, 63-65, 70-76, 78, 79, 81-84, 86-94, 96, 98-103], n=40 |
|  | **Increased equity (qualitative)**<br>**Description:** OCs helped patients communicate with their primary care providers who had previously struggled due to their personal characteristics.<br>**CERQual rating:** High<br>**References:** [32, 41, 42, 44, 47, 63, 70, 71, 74-76, 81-84, 86, 89, 90, 103], n=19 |
| **Patient-centredness**<br>(care that is respectful and responsive) | **Decreased patient satisfaction (qualitative)**<br>**Description:** Negative patient experiences of using OCs.<br>**CERQual rating:** High<br>**References:** [32, 44, 46, 47, 63, 71, 74, 76, 83, 87, 89-91], n=13 |
|  | **Increased patient satisfaction (qualitative and quantitative)**<br>**Description:** Positive patient experiences of using OCs.<br>**CERQual rating:** High<br>**References:** [27, 31, 32, 38, 39, 41, 42, 44, 46, 47, 63, 65, 71, 73, 74, 76, 78, 80-84, 86, 87, 89-93, 95, 97], n=31 |

**Table 3.**
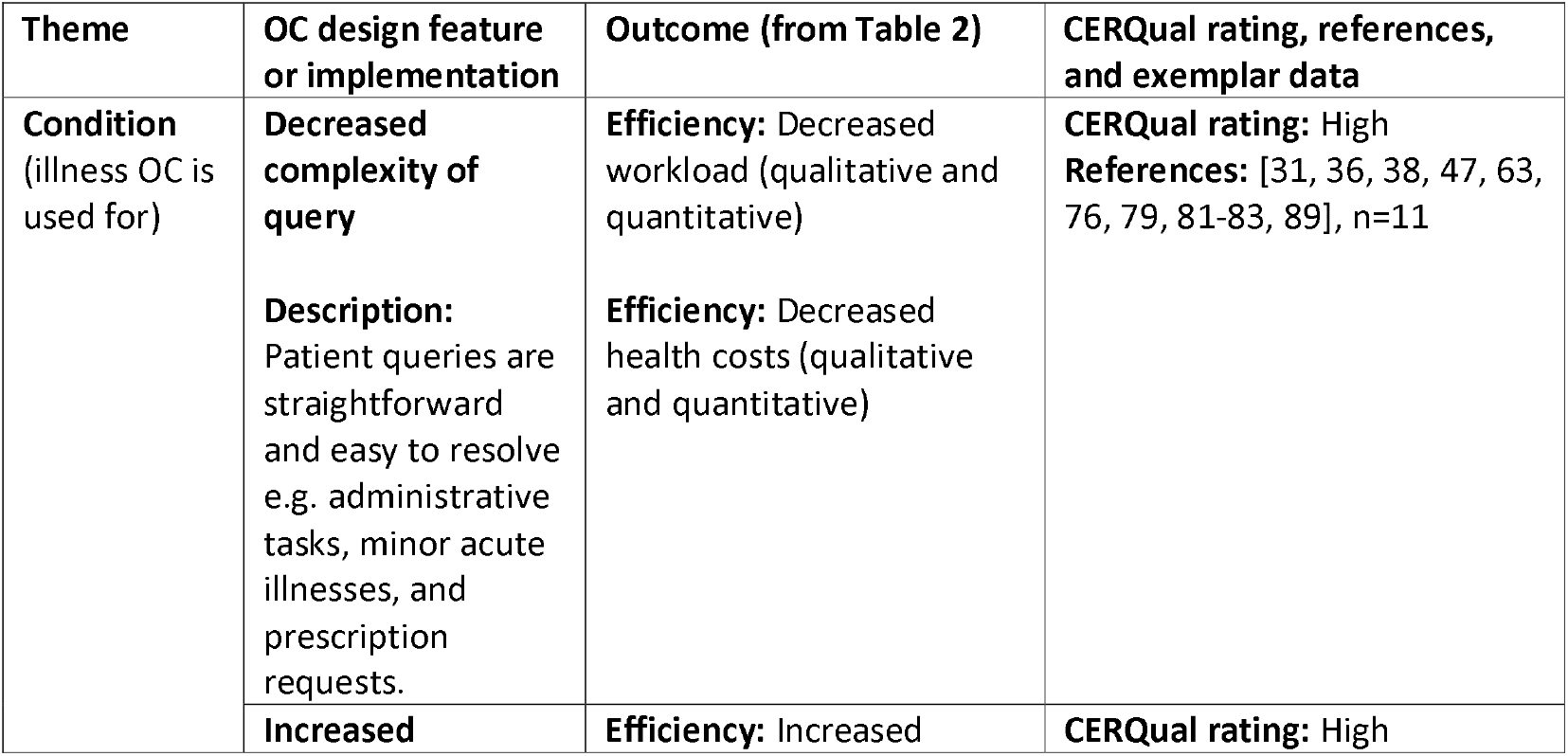
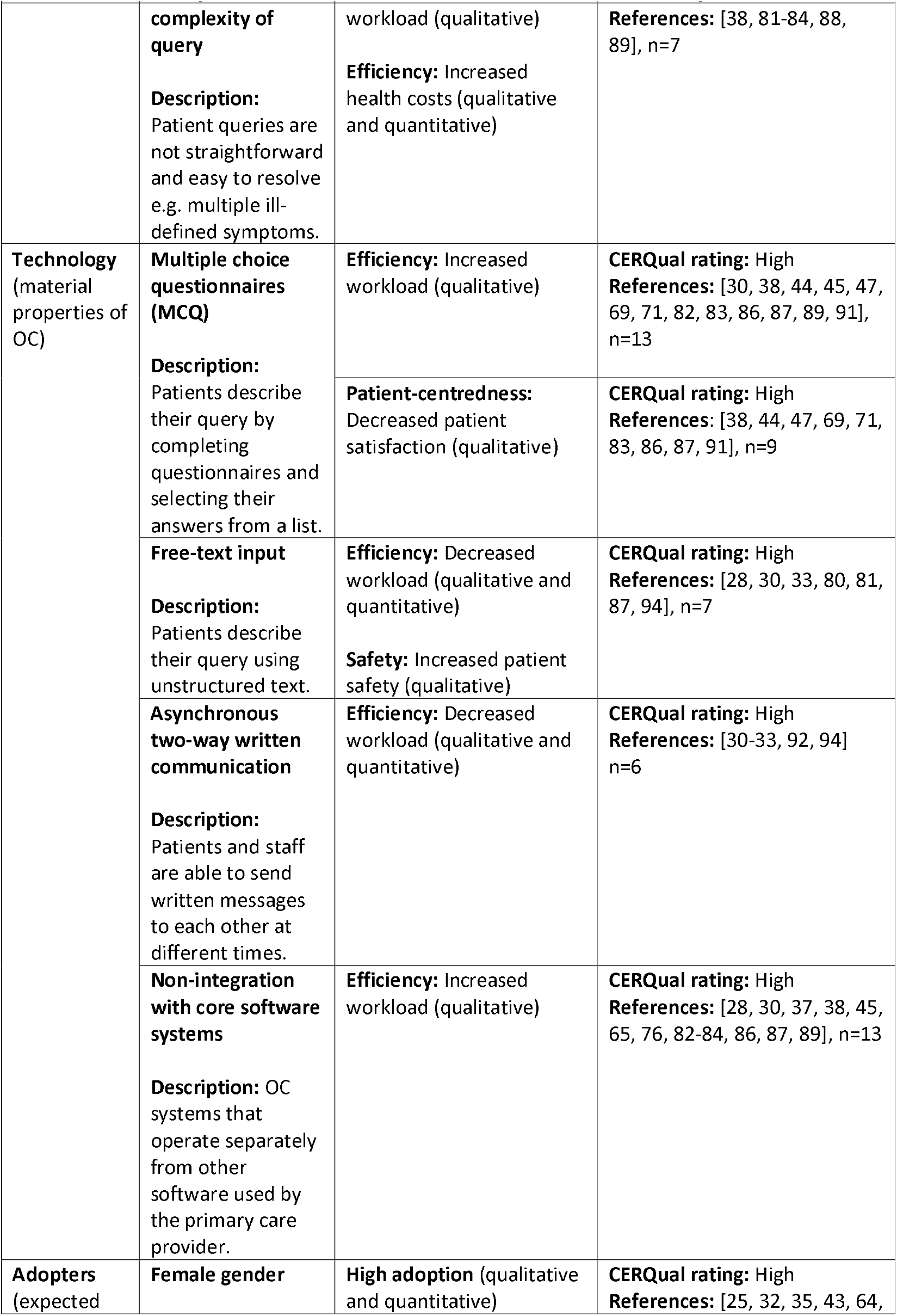
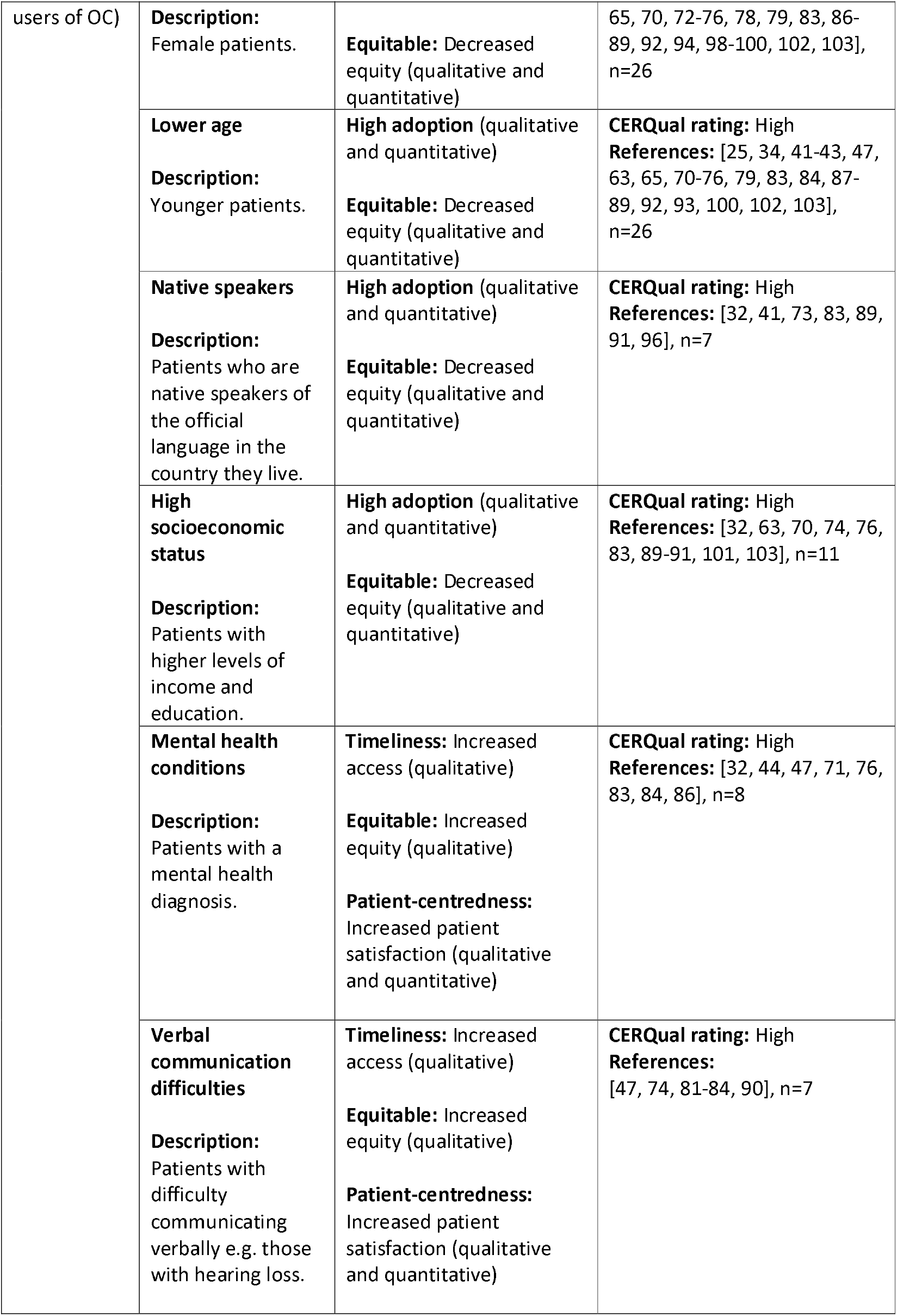
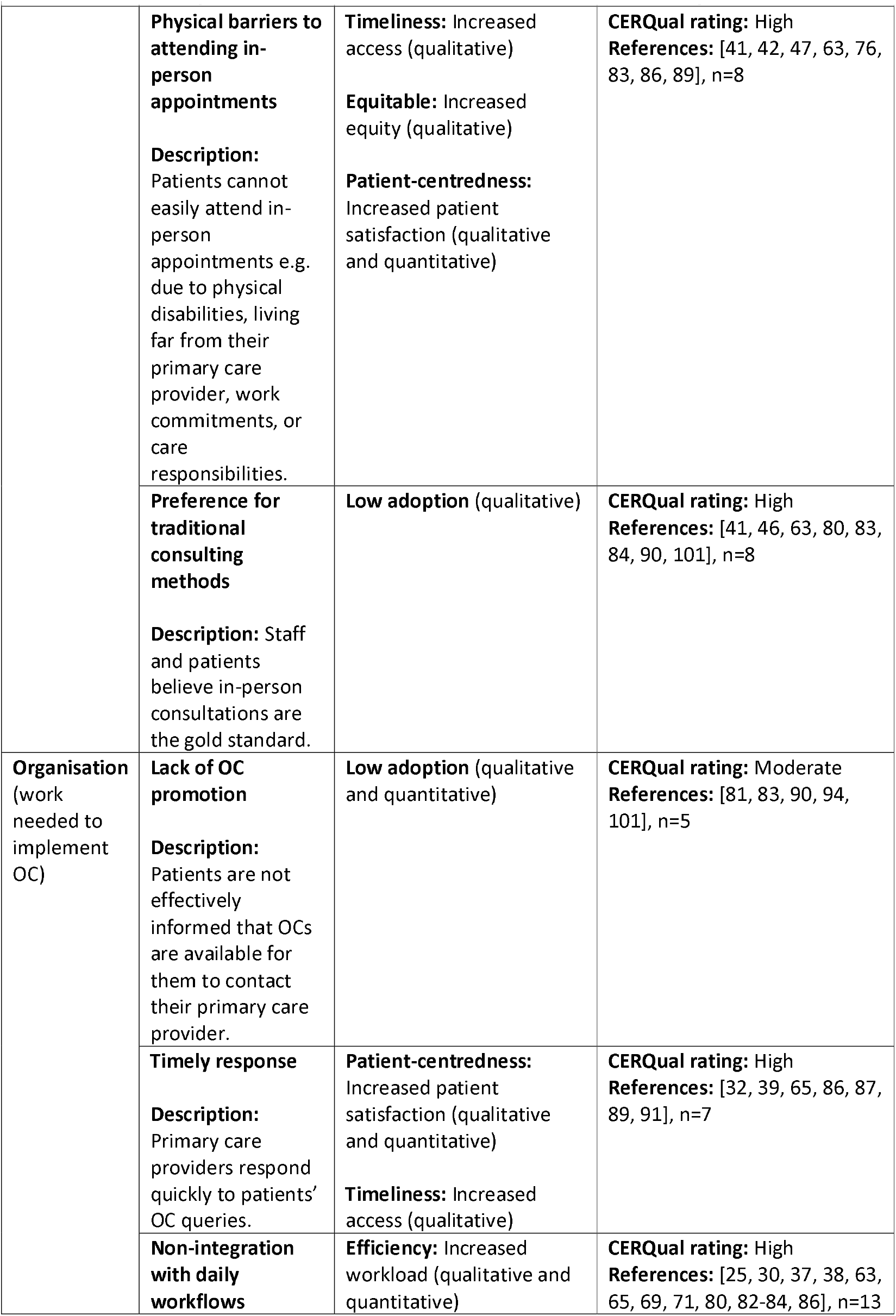
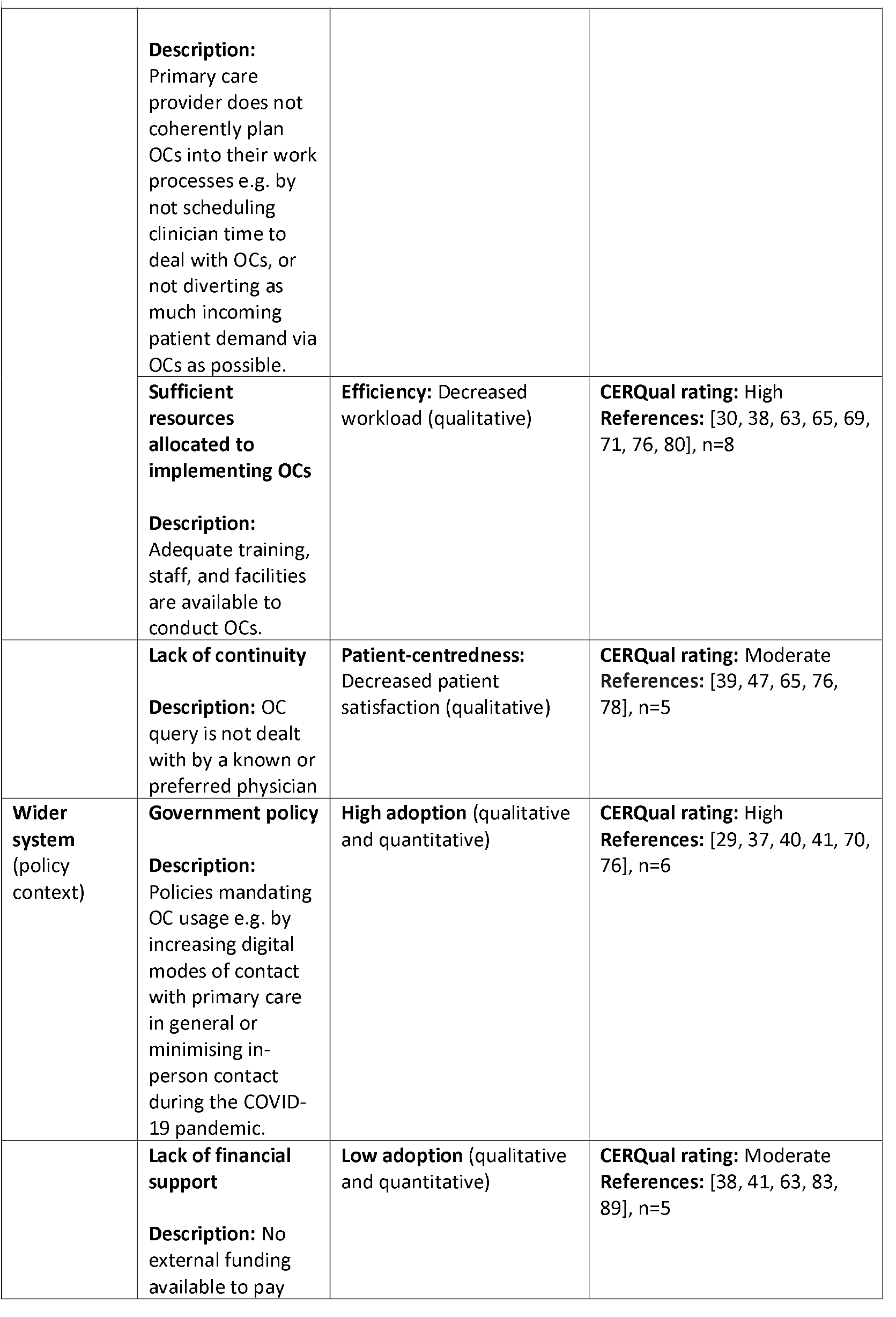
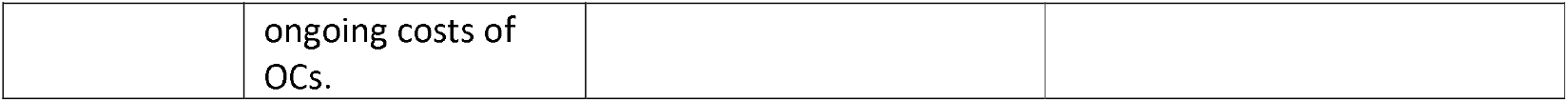
How outcomes of Online Consultations in primary care are influenced by system design and implementation

#### Objective 1: Outcomes of Online Consultations in primary care

##### Safety

In 17 studies, staff and patients expressed general concerns about the impact of OCs on patient safety, particularly around the potential loss of information from patients versus in-person or telephone consultations, and how it could lead to misdiagnosis (e.g. [30]). However, quantitative evidence from 11 studies did not support these concerns in terms of Emergency Department attendance rates (e.g. [78]), hospitalisations (e.g. [79]), deaths (e.g. [72]), and other measures (e.g. [34]). Furthermore, clinicians and patients in 14 studies believed OCs improved patient safety, for example by producing a detailed shared written record of consultations (e.g. [80]) and helping to reduce the spread of communicable diseases such as COVID-19 (e.g. [41]).

##### Timeliness

In 29 studies, OCs were perceived to increase access to primary care services. It was easier and more convenient to make initial contact because patients could submit an OC query at any time without waiting on the phone or attending in-person (e.g. [71]). Once a query was submitted, patients also communicated with health professionals sooner because OCs tended to circumvent the traditional appointment-booking process (e.g. [32]).

##### Efficiency

In total, 33 papers suggested workload decreased for both staff and patients when using OCs. Patient queries were written rather than spoken, incoming phone calls to receptionists reduced (e.g. [81]), and patient histories did not need manual documentation (e.g. [80]). Written queries were usually more detailed than when communicated verbally, and were received by health care staff asynchronously, thus providing opportunities for more objective examination and more effective triage. Consequently, patient queries could more often be directed to other services or dealt with by other staff members, rather than always physicians (e.g. [28]). Combined with their remote nature, OCs also gave staff more autonomy over how their work was organised, thus providing efficiency gains such as working from home and control over how to contact a patient rather than defaulting to an in-person consultation (e.g. [65]). When telephone or in-person consultations were necessary, they were more focused and therefore quicker because the staff member could read the patient query prior to contact (e.g. [82]). OCs reduced workload for patients by avoiding the need to telephone their primary care provider to make an appointment, which often entailed long queues (e.g. [83]), and avoiding in-person consultations when possible, which typically involved travel, waiting rooms, and organising time off work and childcare (e.g. [76]).

In contrast, 29 studies suggested OCs increased workload for staff and patients. Staff described conducting OCs on top of their usual tasks (e.g. [65]) and dealing with them outside normal working hours (e.g. [84]). They believed that because OCs increased access to primary care, patients sought help more readily than they would have previously (e.g. [82]), thus creating ‘supply-induced demand’ [85]. Processing OCs also created new administrative work, such as filing them to EHRs and deciding whether or not they required input from a clinician (e.g. [69]). Workload could also increase for patients if they perceived that entering their query into the OC system was more difficult than explaining them verbally (e.g. [86]).

OCs decreased costs for providers in 20 studies largely by reducing in-person visits, which have associated expenditures related to staffing and utilities (e.g. [87]). Patients reported that due to their convenience, having access to OCs stopped them from visiting other costly unscheduled care providers (e.g. [78]). OCs decreased costs for patients in four studies by avoiding in-person visits, which may entail expenses related to travel, unpaid work leave, and childcare (e.g. [32]).

In contrast, OCs increased costs for providers in nine studies due to associated technology costs (e.g. [41]), time required for clinicians to triage patient queries (e.g. [88]), and insufficient reduction of in-person visits or telephone consultations (e.g. [70]).

##### Equitable

Forty studies suggested that OCs decreased equitable access to care services because their use varied according to patient characteristics (e.g. [41]). Conversely, 19 studies suggested that OCs increased equitable access because they helped particular groups of patients communicate with their primary care providers who had previously struggled (e.g. [71]). These characteristics are discussed in more detail in the Adopters section below.

##### Patient-centredness

Although 13 studies uncovered some patient dissatisfaction with OCs (e.g. [74]), 31 studies found most patients were at least as satisfied with OCs, or were more satisfied with them than traditional in-person appointments (e.g. [27]). Patients liked OCs for the reasons described above: they improved access (Timeliness), reduced their workload and costs (Efficiency), and helped particular groups of patients communicate with their care providers (Equitable).

#### Objective 2: How outcomes of Online Consultations in primary care are influenced by system design and implementation

##### Condition

Eleven studies suggested that OCs decreased staff workload when used for simple queries that were straightforward to resolve because they were more amenable to completion without needing to contact the patient directly via telephone or in-person (e.g. [38]). Simple queries included those related to administrative tasks, new and recurrent minor acute illnesses, prescriptions, tests, requests for advice, follow-up, and some chronic condition reviews (e.g. [31]). These queries also decreased health costs because they saved clinicians time, for example when administrative staff were able to relay messages and there was no direct contact between doctor and patient (e.g. [89]). Seven studies suggested that OCs increased staff workload and costs when used for complex queries, such as those with multiple ill-defined symptoms (e.g. [82]). These queries generally required verbal dialogue with, and physical examination of the patient, and were usually converted to telephone and/or in-person consultations to assess the patient further (e.g. [89]). Staff felt this duplicated the number of contacts with the patient for the same query.

##### Technology

Thirteen studies showed that when patients had to use MCQs to input their OC query it increased both patient and staff workload. Filling out long lists of questions shifted work from the clinician to the patient (e.g. [86]), and staff found them burdensome to read (e.g. [69]). MCQs limited the amount of detail patients could enter so staff could not always fully understand their request. This increased workload because they often had to contact the patient to get further information (e.g. [89]). MCQs also asked questions about seemingly ‘irrelevant’ symptoms, which staff were responsible to assess and follow up, diverting attention away from the patient’s primary concern (e.g. [45]). Due to their restrictive nature, patients regularly adapted their responses to MCQs to get the outcome they wanted, even when it was not the most appropriate use of resources. For example, reporting their symptoms differently to get an in-person consultation when self-care may have been more suitable (‘gaming’; e.g. [82]).

Nine studies suggested that MCQs could also decrease patient satisfaction. Reasons included the amount of work required to complete them (e.g. [71]), their inflexibility in getting the answers patients wanted from their primary care provider (e.g. [44]), and that they could be confusing to navigate (e.g. [91]).

In contrast, seven papers suggested that when patients could primarily report their queries using unstructured free-text, it decreased staff workload and increased patient safety. This was because patients were more able to fully describe their query in sufficient detail using their own words, and clinicians did not have to request further information as often (e.g. [94]).

In six studies, asynchronous two-way written communication within the OC decreased workload for both staff and patients. The ability to reply to patients in writing meant queries could be answered and follow-up questions asked at times convenient to both staff and patients, avoiding lengthy telephone and in-person consultations when appropriate (e.g. [30]). It was also easier to communicate complex information, for example by sending educational materials or using pre-set message templates (e.g. [94]).

Thirteen papers highlighted that a lack of integration between the OC system and other core software used by providers increased staff workload. Non-integration meant staff had to go through multiple steps to perform a task, such as when filing an OC to a patient’s EHR (e.g. [87]).

##### Adopters

Patients using OCs were more likely to be female (n=26, e.g. [79]), younger (n=26, e.g. [75]), a native speaker of the official language in the country they live (n=7, e.g. [91]), and have higher socioeconomic status (n=11, e.g. [32]) than those not using OCs, thus decreasing equity. In contrast, both staff and patients felt that OCs increased access for particular groups of patients who struggled with traditional consultation methods, thus increasing equity and their satisfaction with care. This included patients with: mental health conditions who became anxious when speaking to health professionals on the telephone or in-person (n=8, e.g. [86]); verbal communication difficulties such as hearing loss, who found it easier to communicate in writing (n=7, e.g. [74]); and barriers to attending in-person appointments due to physical disabilities, geography, work commitments, or care responsibilities (n=8, e.g. [89]). Eight papers suggested that when staff and patients viewed traditional in-person methods as the gold standard, it could lead to resistance in adopting OCs (e.g. [84]).

##### Organisation

Five papers found that when OCs were minimally advertised to patients, it led to low rates of adoption (e.g. [90]). Seven papers also showed that responding to a patient’s initial OC query quickly led to high patient satisfaction because it provided an advantage over traditional methods of primary care contact (e.g. [39]); by definition this also increased primary care access.

Thirteen papers found that staff workload was increased when providers did not integrate OCs into their normal daily workflows. For example, not scheduling time for clinicians to deal with OCs meant they were done in addition their normal tasks (e.g. [80]), and not diverting all incoming patient demand via the OC meant different communication routes were often used for the same issue thereby duplicating work (e.g. [38]). Eight papers suggested that provider workload decreased if sufficient resources were allocated to implementing OCs. This included their initial set-up, for example training to enable staff to more effectively handle OCs (e.g. [76]), and their ongoing processing, for example dedicated facilities such as quiet rooms to help staff respond to OCs without distraction (e.g. [30]).

Five papers showed that a lack of continuity between patients and their known doctor impacted negatively on patient satisfaction. This occurred when any doctor could reply to an OC query and patients were not able to specify a doctor to whom to address their query (e.g. [47]).

##### Wider system

Six papers showed that government policies mandating OC usage increased their adoption. Example policies aimed to increase digital modes of contact with primary care in general (e.g. [70]), and to minimise in-person contact during the COVID-19 pandemic (e.g. [41]). Five papers demonstrated that a lack of long-term external financial support for OCs limited their sustainability, as healthcare organisations could often not afford to pay their ongoing costs (e.g. [89]).

## Discussion

### Summary of evidence

We synthesised qualitative and quantitative evidence from 62 studies in nine countries covering 30 OC systems described in detail, with wide-ranging functionalities including AI. Twenty-six studies were published 2020 onwards and 11 were post COVID-19. Overall adoption of OCs by patients was generally low.

Although concerns were raised about patient safety when using OCs, there was no quantitative evidence to support them. Some clinicians and patients believed OCs improved safety, particularly when patients could describe their queries using unstructured free-text. Staff workload decreased when sufficient resources were allocated to implement OCs, and asynchronous two-way written messages to patients could be sent. Workload also decreased when patients used OCs for simple problems or could fully describe their queries using free-text. Staff workload increased when OCs were not integrated with other software or organisational workflows, and when patients used them for complex queries that ultimately required telephone or in-person consultations. OC systems that required patients to describe their queries using multiple choice questionnaires (MCQs) increased workload for both them and staff due to their inflexibility and often irrelevant questions. Health costs were reduced when patients used OCs for simple queries, and increased when used for complex ones.

Patients using OCs were more likely to be female, younger, native speakers, with higher socioeconomic status than those not using OCs, thus decreasing equity. However, OCs increased primary care access for patients who previously struggled with traditional consultation methods, such as those with mental health conditions, verbal communication difficulties, and barriers to attending in-person appointments, thus increasing equity. Access was further increased by providing a timely response to patients’ queries. Patient satisfaction increased when using OCs due to better primary care access, though could decrease when using MCQ formats. Patient satisfaction also decreased when OCs did not support continuity of care. OCs were less likely to be adopted if they were not promoted to patients, if prospective users viewed traditional in-person methods as the gold standard, and if there was a lack of long-term external financial support. Government policies mandating OC usage increased adoption.

### Comparison of findings with other reviews

Consistent with previous reviews relevant to OCs, we found a limited demographic of patients using OCs leading to potential inequitable care [18, 19]. We also found that studies often did not sufficiently explore patients’ perspectives of OCs in-depth [19]; only nine studies (15%) used interview-based methods and had an average sample size of 23. This hampered efforts to understand how such inequities arose.

Contrary to previous reviews we found that OC outcomes are more complex and nuanced than previously reported [18-20]. For example, we identified mixed findings on their impact on workload, patient satisfaction, and equitable care. This contrasts with previous reviews where OCs only increased [20] or had no impact [18] on workload, decreased patient safety [18, 20], and increased inequity [18-20].

These new findings for OCs may be partly explained because 47 (76%) of our included studies had not been covered by these prior reviews. Although there was some overlap of papers (7/57 [18], 7/13 [19], 4/17 [20]), the majority did not meet our inclusion criteria as they were either non-empirical (4/57 [18], 4/13 [19], 4/17 [20]), pre-2010 (26/57 [18], 2/17 [20]), not based in real-world primary care (16/57 [18], 1/13 [19], 6/17 [20]), or did not meet our functional definition of an OC (39/57 [18], 2/13 [19], 6/17 [20]; e.g. symptom checkers with no link to a health professional [104]).

By focusing on design and implementation we have identified new ways in which OCs influence primary care outcomes. For example, we found that by increasing access OCs can increase staff workload by creating ‘supply-induced demand’ [85] (e.g. [82]), and that they can decrease workload by enabling more focused consultations (e.g. [82]). Furthermore, because previous reviews often did not analyse the design or implementation of OCs [18-20], we have identified influential factors that have not previously been described. For example, whilst some reviews identified increased workload when clinicians received insufficient patient information via an OC system (e.g. [19]) we found this was particularly associated with MCQ-based OCs (e.g. [89]). We also identified that allowing patients to describe their queries using unstructured free-text had the opposite effect (e.g. [94]), whilst also having a positive impact on patient safety (e.g. [30]).

### Strengths and limitations

As evidenced by the range of examples in Appendix 1, we adopted a fundamental functional definition of OCs rather than relying on the name given to them by authors of included studies. When combined with our comprehensive searches across multiple databases and inclusion of grey literature, we have identified more empirical studies relevant to OCs than any previous evidence synthesis on the topic [18-20]. Combined with our focus on causal mechanisms, this has helped us develop a new and theoretically informed understanding of OCs that has not previously been reported.

As in all systematic reviews, our synthesis is reliant on what study authors reported. OC features were not always described in sufficient detail to understand how they impacted outcomes (e.g. [40]). There was also a lack of patient perspective in the studies, particularly from OC non-users (e.g. [37]). We made our literature search strategy as inclusive as possible regarding different terms used for OCs (Appendix 1), though due to their wide-ranging nature it is possible that some papers were missed. We updated our searches in November 2021 in order to capture more recently published studies, although due to time constraints only one author (SD) screened these newer papers. This enabled us to capture studies conducted in the context of COVID-19 (n=11, 18% of all included studies).

### Implications for practice and research

Our findings show that OC outcomes are complex, and can be influenced by subtle ways in which they are designed and implemented. To maximise their benefit for patients and staff, we therefore provide recommendations for: OC developers on how systems could be designed; healthcare organisations for how they can be implemented and used; and researchers on questions and areas for further investigation. They are discussed below under the high-level themes from objective 2, and summarised in Table 4.

**Table 4:**
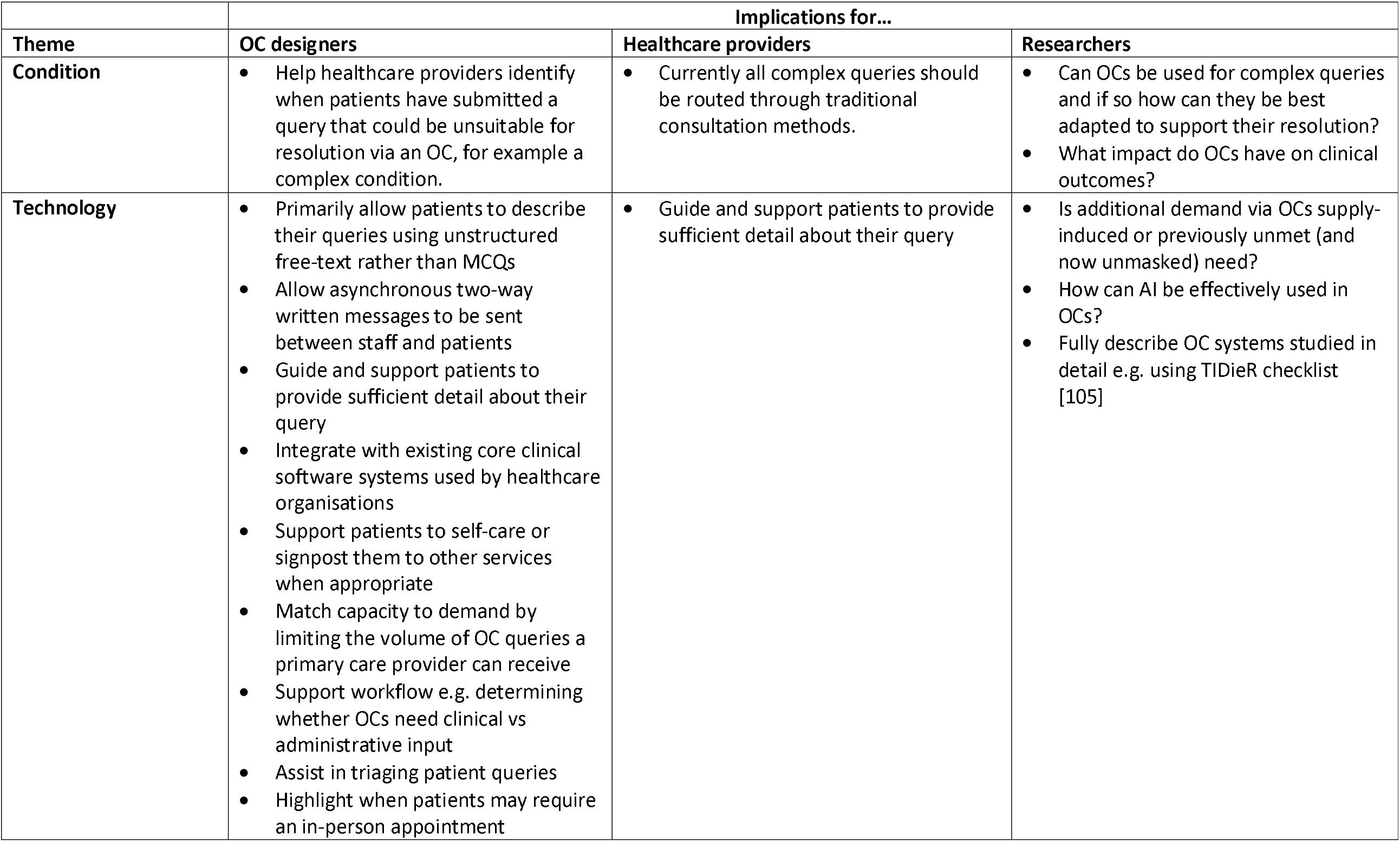
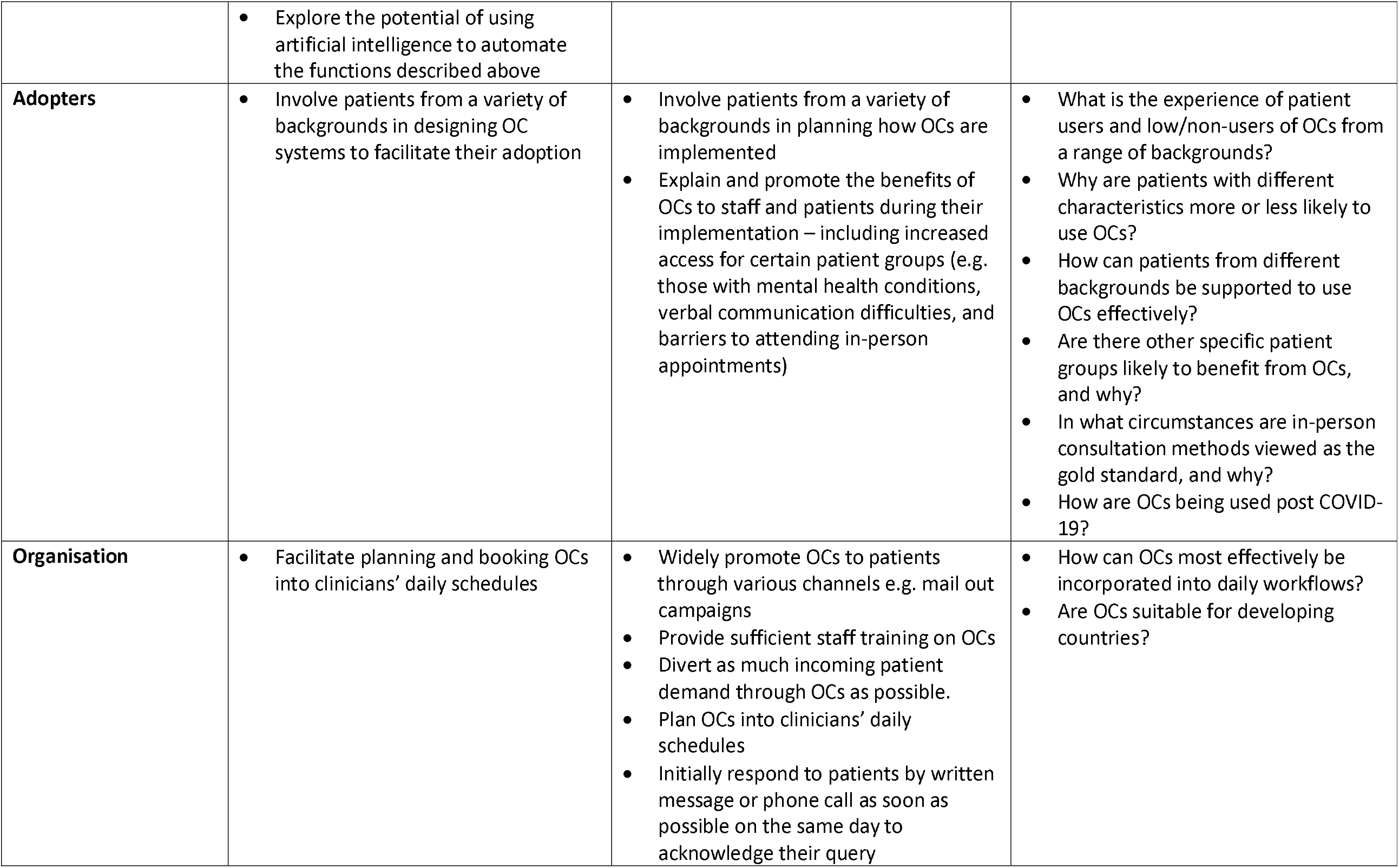
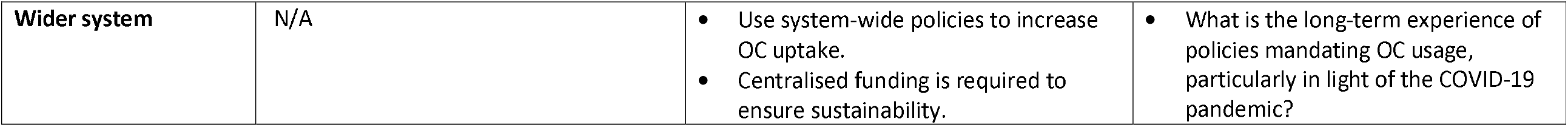
Implications for Online Consultation research and practice

| Theme | Implications for... |  |  |
| --- | --- | --- | --- |
|  | OC designers | Healthcare providers | Researchers |
| **Condition** | Help healthcare providers identify when patients have submitted a query that could be unsuitable for resolution via an OC, for example a complex condition. | Currently all complex queries should be routed through traditional consultation methods. | Can OCs be used for complex queries and if so how can they be best adapted to support their resolution?<br>What impact do OCs have on clinical outcomes? |
| **Technology** | Primarily allow patients to describe their queries using unstructured free-text rather than MCQs<br>Allow asynchronous two-way written messages to be sent between staff and patients<br>Guide and support patients to provide sufficient detail about their query<br>Integrate with existing core clinical software systems used by healthcare organisations<br>Support patients to self-care or signpost them to other services when appropriate<br>Match capacity to demand by limiting the volume of OC queries a primary care provider can receive<br>Support workflow e.g. determining whether OCs need clinical vs administrative input<br>Assist in triaging patient queries<br>Highlight when patients may require an in-person appointment | Guide and support patients to provide sufficient detail about their query | Is additional demand via OCs supply-induced or previously unmet (and now unmasked) need?<br>How can AI be effectively used in OCs?<br>Fully describe OC systems studied in detail e.g. using TIDieR checklist [105] |
|  | Explore the potential of using artificial intelligence to automate the functions described above |  |  |
| **Adopters** | Involve patients from a variety of backgrounds in designing OC systems to facilitate their adoption | Involve patients from a variety of backgrounds in planning how OCs are implemented<br>Explain and promote the benefits of OCs to staff and patients during their implementation – including increased access for certain patient groups (e.g. those with mental health conditions, verbal communication difficulties, and barriers to attending in-person appointments) | What is the experience of patient users and low/non-users of OCs from a range of backgrounds?<br>Why are patients with different characteristics more or less likely to use OCs?<br>How can patients from different backgrounds be supported to use OCs effectively?<br>Are there other specific patient groups likely to benefit from OCs, and why?<br>In what circumstances are in-person consultation methods viewed as the gold standard, and why?<br>How are OCs being used post COVID-19? |
| **Organisation** | Facilitate planning and booking OCs into clinicians' daily schedules | Widely promote OCs to patients through various channels e.g. mail out campaigns<br>Provide sufficient staff training on OCs<br>Divert as much incoming patient demand through OCs as possible.<br>Plan OCs into clinicians' daily schedules<br>Initially respond to patients by written message or phone call as soon as possible on the same day to acknowledge their query | How can OCs most effectively be incorporated into daily workflows?<br>Are OCs suitable for developing countries? |
| **Wider system** | N/A | • Use system-wide policies to increase OC uptake.<br>• Centralised funding is required to ensure sustainability. | • What is the long-term experience of policies mandating OC usage, particularly in light of the COVID-19 pandemic? |

#### Condition

We found that OCs worked more efficiently when used for simple rather than complex patient queries. It is unclear whether OCs are unsuitable for these conditions, or whether workflows and procedures can be better organised, and OC systems better designed to deal with them. As it currently stands, we recommend that complex conditions should be routed through traditional consultation methods (e.g. in-person, telephone), and further research conducted into how these types of conditions could be better handled via OCs to ensure they benefit all patients. One way in which OC designers could help is by highlighting to healthcare providers when patient queries may be unsuitable for resolution via an OC, so they can be dealt with appropriately.

Using the IOM care quality domains to synthesise OC outcomes highlighted an evidence gap for ‘Effectiveness’, which received only low confidence ratings (Appendix 7). This domain refers to care based on scientific knowledge to determine whether an intervention produces better outcomes than alternatives [59]. Although we found evidence of OC impacts on quantitative measures of patient satisfaction (e.g. [27]) and patient safety (e.g. [72]), it is unknown if OCs impact clinical outcomes, such as delivery of evidence-based treatment, or rates of morbidity, and should be investigated further.

#### Technology

We found OCs that allowed patients to fully describe their queries using unstructured free-text rather than via MCQs decreased workload and increased patient safety (e.g. [94]), whereas MCQs could increase both staff and patient workload (e.g. [45]), and decrease patient satisfaction (e.g. [44]). OC developers should therefore provide this functionality, whilst at the same time guiding and supporting patients to provide sufficient detail for their primary care provider to quickly and safely respond to their query. Similarly, we found that OCs with asynchronous two-way written messages decreased workload (e.g. [30]), whereas those that did not integrate with existing core clinical software systems increased workload (e.g. [87]). Where possible, OC developers should therefore also incorporate these designs.

Technology design also has a role in mitigating some of the undesirable outcomes we identified from using OCs, including increasing workload and costs. Increased workload is particularly important because it can lead to a mismatch between patient demand and healthcare resource, which can in turn threaten patient safety if providers are unable to deal with OCs in an appropriate timeframe. One way this could happen is through increased demand: if there are too many OCs submitted by patients and not enough staff to deal with them (e.g. [30]). Whether this additional demand is supply-induced [85] or previously unmet (and now unmasked) need was unclear from the studies we included (e.g. [76]) and requires further research. Nevertheless, OC systems could help by: supporting patients to self-care or signposting them to other services when appropriate; matching capacity to demand by limiting the number of OC queries primary care providers can receive from patients; supporting workflow, for example by determining whether OCs require clinical input or not to relieve workload on administrators (e.g. [69]); assist in triaging patient queries to reduce associated costs of solely relying on clinicians to triage (e.g. [88]); and highlighting when patients may require an in-person appointment to facilitate direct booking to avoid work duplication (e.g. [89]). According to our definition [60], many of these functions may require a form of AI to be most effective, which should be explored by OC designers (Figure 2). Thirteen MCQ-based OC systems in our review used AI (Table 1, e.g. [29]), though mainly for adapting the questions they asked patients during query submission, rather than the functions described above. Furthermore, AI was usually not the focus of the studies, and we consequently found only low confidence evidence regarding its use in OCs (Appendix 8). Therefore, how AI could be used by OC systems in clinical practice requires further research.

Included papers did not always adequately describe the OC systems they studied, limiting our ability to determine how specific features influenced observed outcomes. Future research should describe OC systems in detail so that evaluation findings can be usefully compared, for example by using the template for intervention description and replication (TIDieR) checklist [105].

#### Adopters

OCs offer advantages such as convenience and access to timely care to some patient groups, but for the technology to be equitable it must benefit all patients. Similar to related reviews [18, 19], we found patient characteristics (age, gender, native language, and socioeconomic status) to be important factors in OC adoption. Qualitative evidence contributed to these findings (e.g. [89]), however as described above, there was insufficient exploration of participant (especially patient) experiences to confidently explain why inequitable care arose, or how it could be overcome. Study authors and healthcare staff speculated reasons such as older patients may be less adept at using modern technology (e.g. [83]), however we were unable to formulate evidence-based hypotheses. Future research should therefore explore perspectives from patients using (and not using) OCs from a wide range of backgrounds using in-depth qualitative techniques such as interview-based methods. Patients from a variety of backgrounds should also be involved in how OC systems are designed, and help plan how they are implemented in practice.

In contrast, we found that OCs increased access for patients who traditionally struggled to access primary care, such as those with mental health conditions and verbal communication difficulties. These should be viewed as benefits and promoted as such to health care providers and patients. Further research should explore other patient groups that could specifically benefit from OCs.

Staff and patients resisted adopting OCs when they viewed traditional in-person consultation methods as the gold standard. Although this was understandable for complex queries (e.g. [82]), it was unclear whether other factors also influenced this view. Future research should address this evidence gap, particularly since COVID-19 has made remote consultations more commonplace [22]. In the meantime, this perception could be challenged by explaining the benefits of OCs found in our review to prospective users [106].

#### Organisation

In order for patients and staff to experience the benefits of OCs, they must be widely promoted to patients as a route for them to contact their primary care provider. This can happen through various channels, such as mail out campaigns (e.g. via SMS) or by verbally mentioning OCs when in contact with patients (e.g. when receptionists speak to patients on the telephone).

To minimise workload associated with OCs, organisations should allocate sufficient resources to both setting up and processing them. This includes providing adequate training on how to use OCs, and ensuring there are enough staff and facilities (e.g. computers and rooms) to deal with them. They should integrate OCs into their daily workflows by diverting as much incoming patient demand as possible through the system (e.g. by advising patients who telephone to use the OC system instead) and scheduling time for staff to deal with them. Having one communication channel avoids duplication and increases the proportion of patient contacts that benefit from the advantages of OCs. Traditional ‘appointments’ or slots in clinicians’ schedules could be reserved for OCs to give them adequate time to respond and ensure they do not become additional tasks to complete on top of their normal work. This has the added benefit of reducing costs by replacing other more expensive forms of consultation, such as in-person appointments. OC system developers can help by providing staff and patients the functionality to ‘book’ into these schedules automatically. We found little exploration of this approach in our included studies, and it therefore requires further research as to how it could be implemented most effectively.

Our findings show that providers can increase access and patient satisfaction by responding quickly to OCs, though definitions of what this involved were unclear. We recommend providing an initial response to patients’ OC queries as soon as possible on the same day – either by asynchronous written message or telephone call. This does not mean the entire query needs to be resolved at this point, just that initial contact has been made and the query acknowledged. The OC can then be booked into a clinician’s schedule (if necessary) to fully deal with it as described above, whilst considering competing clinical priorities and available capacity.

We included studies from nine countries, all of which were developed western countries. Due to their remote nature, OCs may have a role in developing countries where there are isolated communities and fewer health care staff per head of population. However, further research is required to understand how their technological and financial barriers could be overcome.

#### Wider system

Governmental policies to promote OCs are effective at increasing adoption, though centralised funding is needed to sustain their use. It is unclear what the long-term experience of such policies are from the papers we included, particularly in response to those relating to the COVID-19 pandemic.

## Conclusions

This is the first theoretically-informed synthesis of empirical research on OCs in primary care, and uniquely includes studies conducted during COVID-19. It contributes new knowledge that OCs are safe and can have positive outcomes, such as increased access to primary care and decreased patient costs. However, they are also complex and often produce conflicting outcomes related to provider costs, workload, patient satisfaction, and equitable care. Some of these are unintended, and conflict with the promotion of OCs by policymakers as a way to address already increasing workload and decreasing workforce capacity in primary care [4-9]. Unlike previous evidence syntheses on the topic, we have shown that negative OC outcomes can be mitigated by appropriate OC system design (e.g. free-text vs MCQ formats, asynchronous two-way communication), incorporating advanced technologies (e.g. AI), and integration into technical (e.g. software) and organisational (e.g. timely responses) workflows. Since the advent of COVID-19, OCs have become indispensable, though further engineering and implementation research is required to realise their full benefits.

## Registration and protocol

The study protocol was registered with PROSPERO (CRD42020191802) [107]. The original title was amended to be less general and more specific to the objectives of the review. Objective 2 was amended to include all identified outcomes, rather than limited to the ones originally specified.

## Data Availability

All papers reviewed in our systematic review are
publicly available.

## Acknowledgements

This research was funded by Innovate UK [105178] and a Wellcome Trust Clinical Research Career Development Fellowship for BCB [209593/Z/17/Z]. NP’s time was partially funded by the National Institute for Health Research Greater Manchester Patient Safety Translational Research Centre (NIHR Greater Manchester PSTRC). The views expressed are those of the authors and not necessarily those of the NHS, the NIHR, or the Department of Health and Social Care. NIHR had no role in study design, data collection, data analysis, data interpretation, or writing of the report. The corresponding author had full access to all of the data and the final responsibility to submit for publication.

SD, TC, and BCB refined the research question; developed the search strings; conducted screening, critical appraisal, data extraction and data analysis; and wrote the first draft of the manuscript. NP contributed to the conception and design of the review. All authors contributed to the final analysis, and approved the final submitted version of the manuscript.

## Abbreviations

AI: Artificial Intelligence
EHR: Electronic Health Record
GRADE-CERQual: Grading of Recommendations Assessment, Development, and Evaluation - Confidence in the Evidence from Reviews of Qualitative research
IOM: Institute of Medicine
MMAT: Mixed Methods Appraisal Tool
NASSS: Non-adoption, abandonment, scale-up, spread, sustainability (framework)
MCQ: Multiple choice questionnaire
NHS: National Health Service
OCs: Online Triage and Consultation systems
PRISMA: Preferred Reporting Items for Systematic Reviews and Meta-Analyses
PROSPERO: The International Prospective Register of Systematic Reviews
UK: United Kingdom
US: United States

