## Supplementary material for "Understanding how the design and implementation of Online Consultations influence primary care outcomes: Systematic review of evidence with recommendations for designers, providers, and researchers": Figure 1

### Slide 1
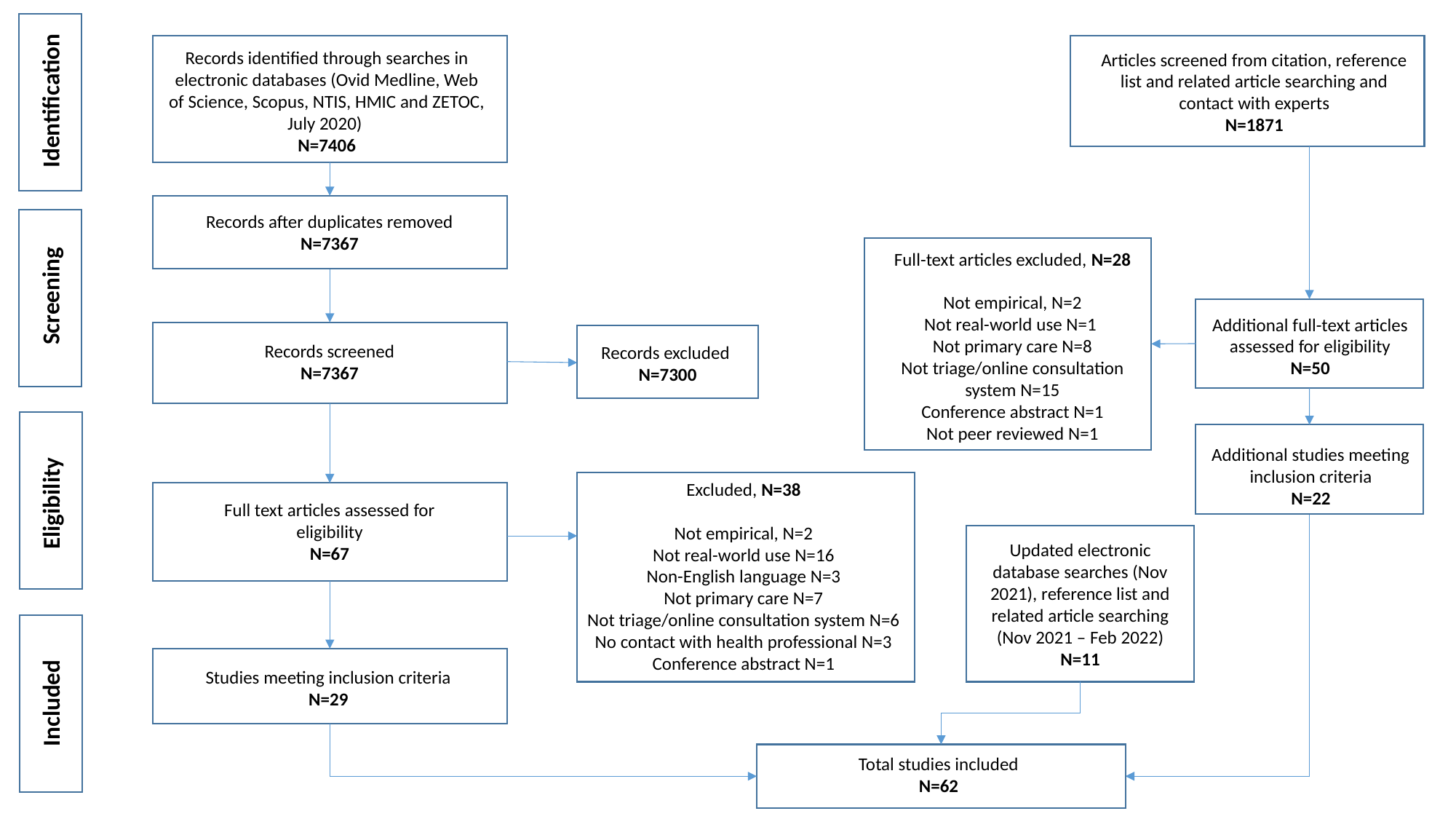

Records identified through searches in electronic databases (Ovid Medline, Web of Science, Scopus, NTIS, HMIC and ZETOC, July 2020)
N=7406
Articles screened from citation, reference list and related article searching and contact with experts
N=1871
Identification
Records after duplicates removed
N=7367
Full-text articles excluded, N=28
Not empirical, N=2
Not real-world use N=1
Not primary care N=8
Not triage/online consultation system N=15
Conference abstract N=1
Not peer reviewed N=1
Screening
Additional full-text articles assessed for eligibility
N=50
Records screened
N=7367
Records excluded
N=7300
Additional studies meeting inclusion criteria
N=22
Excluded, N=38
Not empirical, N=2
Not real-world use N=16
Non-English language N=3
Not primary care N=7
Not triage/online consultation system N=6
No contact with health professional N=3
Conference abstract N=1
Eligibility
Full text articles assessed for eligibility
N=67
Updated electronic database searches (Nov 2021), reference list and related article searching (Nov 2021 – Feb 2022)
N=11
Studies meeting inclusion criteria
N=29
Included
Total studies included
N=62
