## Supplementary material for "Understanding how the design and implementation of Online Consultations influence primary care outcomes: Systematic review of evidence with recommendations for designers, providers, and researchers": Figure 2

### Slide 1
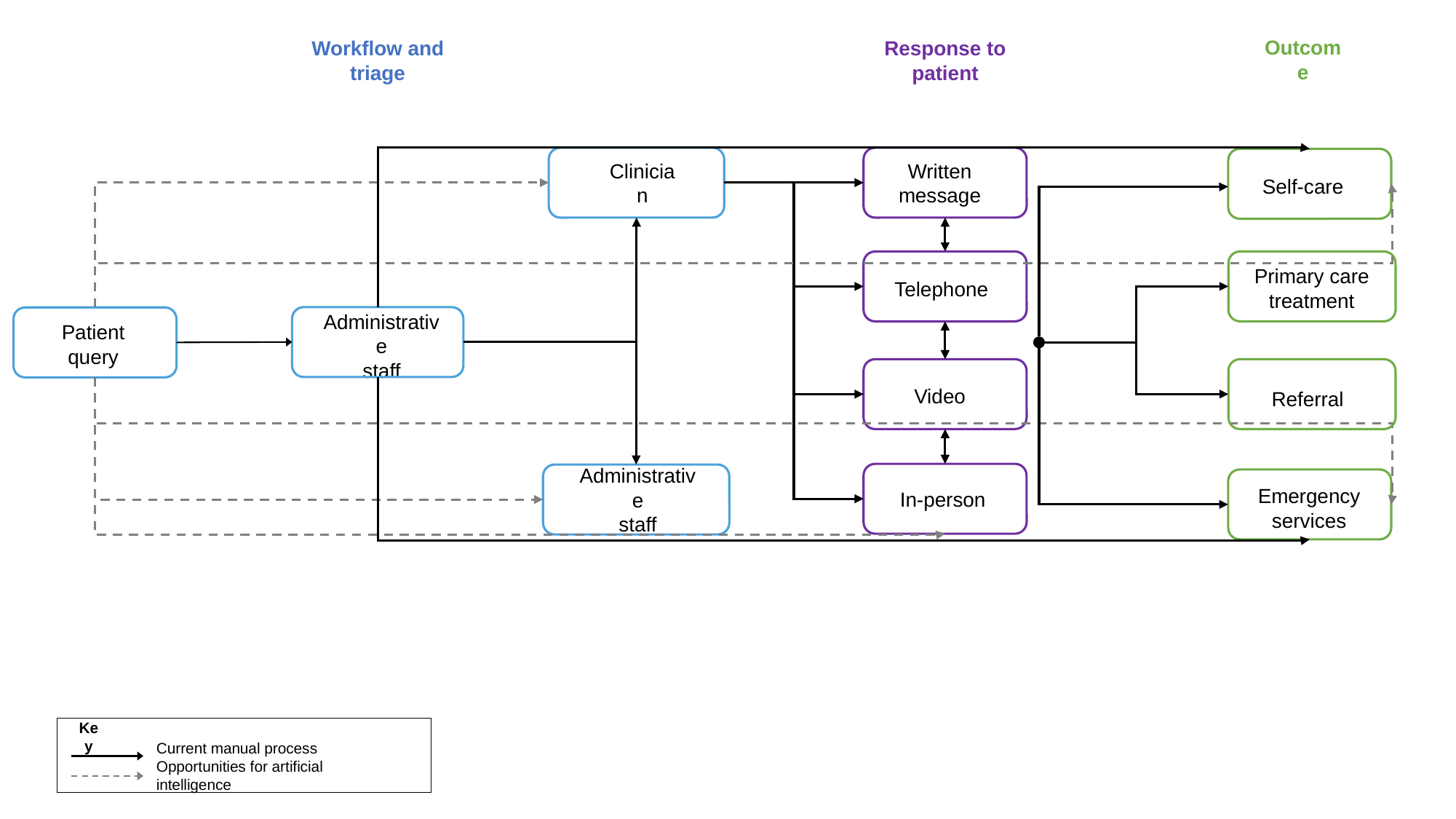

Outcome
Response to patient
Workflow and triage
Written message
Clinician
Self-care
Primary care treatment
Telephone
Administrative
staff
Patient query
Video
Referral
Administrative
staff
Emergency services
In-person
Key
Current manual process
Opportunities for artificial intelligence
