## Appendix 1 for "Understanding how the design and implementation of Online Consultations influence primary care outcomes: Systematic review of evidence with recommendations for designers, providers, and researchers"

**Appendix 1: Terms used by included studies for Online Consultations**

| **Term** | **Papers** |
| --- | --- |
| Triage system based on a symptom-checker | [1] |
| Asynchronous chat | [2] |
| Digital Care | [3] |
| Digital health consultations | [4] |
| Digital healthcare platform | [5] |
| Digital Primary Health Care (DPHC) | [6-8] |
| Digital Doctor Reception | [9] |
| Digital communication system | [10] |
| Digital chat-based communication system | [11] |
| Electronic consultations/e-consultation/e-consult | [12-27] |
| eVisit/e-Visit | [28-47] |
| Online consultation | [48-51] |
| Online visit | [52] |
| Online triage | [53] |
| Self-triage/self-scheduling | [54] |
| Teleconsultation | [55-57] |
| Virtual consulting | [58] |
| Virtual visit | [59-62] |

7.	Health Innovation Manchester. GM Digital First Primary Care: Patient and public insights: Workshop results. Report. Manchester: Health Innovation Manchester, 2021 Sept 2021. Report No.: 1 Contract No.: 1 Sept.
