## Appendix 2 for "Understanding how the design and implementation of Online Consultations influence primary care outcomes: Systematic review of evidence with recommendations for designers, providers, and researchers"

**Appendix 2: Search terms**

| **Database searched** | **Date of Search** | **Search Terms** | **Filters / Limiters applied** |
| --- | --- | --- | --- |
| Ovid Medline | 30/06/2020 | S1: triage.mp. or Triage/; S2: e-triage; S3: Diagnostic Self Evaluation/ or Self-triage.mp; S4: 1 OR 2 OR 3; S5: consultation.mp; S6: Remote Consultation/ or e-consultation.mp.; S7: S5 OR S6; S8: technology.mp. or Technology/ or Information Technology; S9: electronic.mp. or Electronics/; S10: Digital.mp.; S11: Online.mp.; S12: Mobile Applications/; S13: 8 or 9 or 10 or 11 or 12; S14: primary care.mp. or Primary Health Care/; S15: general practice.mp. or General Practice/; S16: family practice.mp. or Family Practice/; S17: 14 OR 15 OR 16; S18: S4 AND S13; S19: S7 AND S13; S20: S18 AND S17; S21: S19 AND S17; S22: S20 OR S21 | 2010 - CURRENT |
| Ovid Embase | 14/07/2020 | S1: triage.mp. or Triage/; S2: e-triage; S3: Diagnostic Self Evaluation/ or Self-triage.mp; S4: 1 OR 2 OR 3; S5: consultation.mp; S6: Remote Consultation/ or e-consultation.mp.; S7: S5 OR S6; S8: technology.mp. or Technology/ or Information Technology; S9: electronic.mp. or Electronics/; S10: Digital.mp.; S11: Online.mp.; S12: Mobile Applications/; S13: 8 or 9 or 10 or 11 or 12; S14: primary care.mp. or Primary Health Care/; S15: general practice.mp. or General Practice/; S16: family practice.mp. or Family Practice/; S17: 14 OR 15 OR 16; S18: S4 AND S13; S19: S7 AND S13; S20: S18 AND S17; S21: S19 AND S17; S22: S20 OR S21 | 2010 - CURRENT |
| Web of Science (Core Collection) | 21/07/2020 | S1: triage; S2: e-triage; S3: self-triage; S4: "Diagnostic Self Evaluation"; S5: S1 OR S2 OR S3 OR S4; S6: Consultation; S7: "Remote Consultation"; S8: E-consult*; S9: S6 OR S7 OR S8; S10: Technolog*; S11: "Information Technolog*"; S12: Electronic*; S13: Digital; S14: Online; S15: Mobile Phone app*; S16: S10 OR S11 OR S12 OR S13 OR S14 OR S15; S17: "Primary care"; S18: "Primary health care"; S19: "General practi*"; S20: "Family practi*"; S21: S17 OR S18 OR S19 OR S20; S22: S5 AND S16; S23: S9 AND S16; S24: S22 AND S21; S25: S23 AND S21; S26: S24 OR S25 | 2010 - CURRENT |
| Scopus | 23/07/2020 | S1: triage; S2: e-triage; S3: self-triage; S4: "Diagnostic Self Evaluation"; S5: S1 OR S2 OR S3 OR S4; S6: Consultation; S7: "Remote Consultation"; S8: E-consult*; S9: S6 OR S7 OR S8; S10: Technolog*; S11: "Information Technolog*"; S12: Electronic*; S13: Digital; S14: Online; S15: Mobile Phone app*; S16: S10 OR S11 OR S12 OR S13 OR S14 OR S15; S17: "Primary care"; S18: "Primary health care"; S19: "General practi*"; S20: "Family practi*"; S21: S17 OR S18 OR S19 OR S20; S22: S5 AND S16; S23: S9 AND S16; S24: S22 AND S21; S25: S23 AND S21; S26: S24 OR S25 | 2010 - CURRENT |
