## Appendix 3 for "Understanding how the design and implementation of Online Consultations influence primary care outcomes: Systematic review of evidence with recommendations for designers, providers, and researchers"

**Appendix 3: Data extraction form**

**STUDY DETAILS**

| **Study ID** |
| --- |
| **Date data extracted** |
| **Study Citation**  (Title, author(s), journal, year) |
| **Research objective(s)/question of study** |
| **Research design and methods** (including sample size and location/s and scale of study) |
| **Main findings** |
| **Outcomes** (primary: qualitative or quantitative outcomes of online triage and consultation systems; secondary: facilitating and inhibiting factors) |
| **Degree of adoption, nonadoption, abandonment, spread, scale-up, and sustainability** (rated as low, medium, high) |
| **Clinical condition** (included or excluded, nature of condition, comorbidities, socio-cultural influences) |
| **Type of technology** (material features, type of data generated, knowledge needed to use, technology supply model) |
| **Value proposition** (supply-side value – to developer, demand-side value – to patient) |
| **Adopters** (staff, patient, carers) |
| **Organisation** (capacity to innovate, readiness for the technology, nature of adoption, extent of change needed to routine, work needed to implement change) |
| **Wider system** (political / policy, regulatory / legal, professional, socio-cultural) |
| **Embedding and adaptation over time** (scope for adaptation over time, organisational resilience) |

**RISK OF BIAS (from the MMAT)**

| **Category of study designs** | **Methodological quality criteria** | **Responses** | | | |
| --- | --- | --- | --- | --- | --- |
|  |  | Yes | No | Can’t tell | Comments |
| Screening questions (for all types) | S1. Are there clear research questions? |  |  |  |  |
|  | S2. Do the collected data allow to address the research questions? |  |  |  |  |
|  | *Further appraisal may not be feasible or appropriate when the answer is ‘No’ or ‘Can’t tell’ to one or both screening questions.* | | | | |
| 1. Qualitative | 1.1. Is the qualitative approach appropriate to answer the research question? |  |  |  |  |
|  | 1.2. Are the qualitative data collection methods adequate to address the research question? |  |  |  |  |
|  | 1.3. Are the findings adequately derived from the data? |  |  |  |  |
|  | 1.4. Is the interpretation of results sufficiently substantiated by data? |  |  |  |  |
|  | 1.5. Is there coherence between qualitative data sources, collection, analysis and interpretation? |  |  |  |  |
| 2. Quantitative randomized controlled trials | 2.1. Is randomization appropriately performed? |  |  |  |  |
|  | 2.2. Are the groups comparable at baseline? |  |  |  |  |
|  | 2.3. Are there complete outcome data? |  |  |  |  |
|  | 2.4. Are outcome assessors blinded to the intervention provided? |  |  |  |  |
|  | 2.5 Did the participants adhere to the assigned intervention? |  |  |  |  |
| 3. Quantitative nonrandomized | 3.1. Are the participants representative of the target population? |  |  |  |  |
|  | 3.2. Are measurements appropriate regarding both the outcome and intervention (or exposure)? |  |  |  |  |
|  | 3.3. Are there complete outcome data? |  |  |  |  |
|  | 3.4. Are the confounders accounted for in the design and analysis? |  |  |  |  |
|  | 3.5. During the study period, is the intervention administered (or exposure occurred) as intended? |  |  |  |  |
| 4. Quantitative descriptive | 4.1. Is the sampling strategy relevant to address the research question? |  |  |  |  |
|  | 4.2. Is the sample representative of the target population? |  |  |  |  |
|  | 4.3. Are the measurements appropriate? |  |  |  |  |
|  | 4.4. Is the risk of nonresponse bias low? |  |  |  |  |
|  | 4.5. Is the statistical analysis appropriate to answer the research question? |  |  |  |  |
| 5. Mixed methods | 5.1. Is there an adequate rationale for using a mixed methods design to address the research question? |  |  |  |  |
|  | 5.2. Are the different components of the study effectively integrated to answer the research question? |  |  |  |  |
|  | 5.3. Are the outputs of the integration of qualitative and quantitative components adequately interpreted? |  |  |  |  |
|  | 5.4. Are divergences and inconsistencies between quantitative and qualitative results adequately addressed? |  |  |  |  |
|  | 5.5. Do the different components of the study adhere to the quality criteria of each tradition of the methods involved? |  |  |  |  |

**MAIN STRENGTHS/LIMITATIONS**

| **Main strengths of study**  (according to reviewer – i.e. you) |
| --- |
| **Main strengths of study**  (according to study author) |
| **Main limitations of study**  (according to reviewer – i.e. you) |
| **Main limitations of study**  (according to study author) |

**SUPPLEMENTARY SEARCHES**

| **Papers in bibliography that may be relevant (out of how many?)**   - Out of how many? - New papers to synthesise or for background discussion |
| --- |
| **Papers in citation searching that may be relevant (using Google Scholar)**   - Out of how many? - New papers to synthesise or for background discussion |
| **Papers in related article searching that may be relevant (Using Google Scholar – limit to first 100 results)**   - Out of how many? - New papers to synthesise or for background discussion |

**MEMOS**
