## Appendix 4 for "Understanding how the design and implementation of Online Consultations influence primary care outcomes: Systematic review of evidence with recommendations for designers, providers, and researchers"

**Appendix 4: Descriptive summary of included studies**

| **Year** | **Publication** | **Authors** | **Research objectives** | **OC system^[1]^** | **Research design** | **Main findings** | **Quality [1]** |
| --- | --- | --- | --- | --- | --- | --- | --- |
| 2022 | British Journal of General Practice | Turner et al. [2] | To identify and understand the unintended  consequences of online consultations in primary  care | Multiple unspecified systems | Qualitative: semi-structured interviews with patients (n=19) and staff (n=18) from 8 General Practices in England, UK | Some patients found it difficult to communicate effectively with a GP,  digitally-excluded patients were disadvantaged. Some staff described additional work, isolation, and dissatisfaction. | 60% |
| 2021 | Workshop results report | Health Innovation Manchester [3] | To gather lived experiences and insights from a diverse group of Greater Manchester patients to inform the digital primary care programme | Digital primary health care - unspecified | Qualitative: Two Patient Engagement  Workshops (n=15 participants), Manchester, UK. | Patients perceived benefits of using digital health but had general and accessibility concerns. | 20% |
| 2021 | Frontiers in Public Health | Landgren and Cajander [4] | To identify reasons for non-use among elderly in the countryside and describe perceived possible challenges and opportunities with digital health consultations. | Digital health consultations - unspecified | Qualitative: Semi-structured interviews were conducted with 13 persons over 65 years old residing in the Swedish countryside. | There was a mistrust for services offered by private companies and their public funding, lack of knowledge of available services, lack of perceived usefulness. Personal interaction and continuity was more important than time or travel conveniences. | 60% |
| 2021 | BMJ Open Quality | Leung and Qureshi [5] | To evaluate the number of high frequency users of Dr iQ and to decrease subsequent monthly usage frequency of all Dr iQ high frequency users | Dr iQ | Mixed methods: Case study (inner city practice, London, UK), two Plan–Do- Study–Act cycles (12 patients and 3 patients) involving interviews, discussion among multidisciplinary team. | The majority of high frequency users had unmet health needs and felt a lack of continuity of care on Dr iQ. They often had complex physical and mental health problems. | 40% |
| 2021 | British Journal of General Practice | Murphy et al. [6] | To investigate the rapid implementation of remote consulting and explore impact over the initial months of the COVID-19 pandemic. | E-consultations – not specified | Mixed methods:  Observational analysis of volume and type of consultations and comparison with previous year and between patient groups.  Staff interviews (n=87)  21 general practices in Bristol, North Somerset, South Gloucestershire, UK | E-consultations registered by a clinician were <1% of all consultations in July 2020. Slow introduction of e-consultations led them to be seen as an additional stream of work.  GPs raised concerns that e-consultations would be used “inappropriately”; would lead to “double doing”; and would enforce existing health inequities. | 20% |
| 2021 | BMC Health Services Research | Nijhof et al. [7] | To explore user experiences and usage patterns of an online consultation system with regard to selected acute and chronic conditions. | e-consultation - unspecified | Quantitative: e-consultation data analysed from 225 patients in the UK for the conditions: tonsillitis, cough, urinary tract infection, acne, vaginosis, sinusitis, and eczema. | Average satisfaction score of 4.15/5. 47.6% of patients had experienced a previous episode of the health condition they were seeking consultation for, 72% had existing comorbidities. Follow-up activity occurred for 87.3% of patients, 66.1% of which included at least one follow-up visit for the same condition as the initial online consultation. | 60% |
| 2021 | Journal of Medical Internet Research | Nilsson et al. [8] | To explore patients’ experiences using a chat-based and automated medical history–taking service in regular, tax-based, not-for-profit primary care in Sweden. | Digital chat-based communication system (unspecified) | Qualitative: Semi-structured interviews (25 patients from 5 primary care centers (PCCs) in Sweden | Asynchronicity allowed patients to take control of the conversation and initiate a chat at any time; however, it could also lead to lengthy conversations which could take days to close. The ability to upload photographs made some visits to the PCC redundant. | 100% |
| 2021 | Telemedicine and e-Health | Penza et al. [9] | To investigate whether e-visits for the management of acute sinusitis have equivalent clinical outcomes for patients when compared with face-to-face visits and nurse administered phone protocols | Secure patient portal, e-visit – not specified | Quantitative: Retrospective chart review. Patients 18-75 years who had a clinical encounter for acute sinusitis. Mayo clinic, US. Comparison of antibiotic prescribing rate, follow-up rates.  150 encounters from each type compared (n=450). | Both e-visit and phone protocol sinusitis encounters were less likely to result in initial treatment with an antibiotic than a face-to-face visit. No significant difference in follow-up rate between e-visits and face-to-face. E-visits had significantly fewer follow-up visits than phone protocol. | 60% |
| 2021 | Journal of the American Board of Family Medicine | Tarn et al. [10] | To examine use of office resources by primary care patients who were initially evaluated through telehealth, telephone, or in-person encounters. | Telehealth: smartphone application to access a patient portal for the visit. | Quantitative: Retrospective electronic health record review of patients (N=202) presenting to a US community-based academic family medicine practice, with COVID-19 symptoms. | Primary care providers used virtual visits to care for most patients presenting with potential COVID-19 symptoms, with many patients choosing telephone over telehealth visits. | 60% |
| 2021 | New Zealand Medical Journal | Wilson et al. [11] | To explore experiences of New Zealand general practice teams in their use of telehealth during the early stages of the COVID-19 pandemic in New Zealand. | Virtual consulting - unspecified | Qualitative: survey of a general practice team members in New Zealand (n=164) | Benefits included rapid triage, convenience and efficiency. Limitations included financial and technical barriers for practices and patients and concerns about clinical risk. | 80% |
| 2020 | Quality and User Experience | Cajander et al. [12] | How has the digitalized and automated patient-centric service affected the work engagement of nurses in primary care? | Chat automation system (unspecified) | Qualitative: Contextual inquiries (n=5) and semi-structured interviews (n=9) with nurses in primary care in Sweden. | Nurses experienced less time pressure and emotional pressure, but also a loss of job control and feedback from colleagues working from home. | 60% |
| 2020 | JMIR Human Factors | Eldh et al. [13] | To describe the experience of health care staff using an OC | Flow | Qualitative: Interviews with health care staff (n=21) in 5 primary care centers in Sweden. | OC implementation is shaped by the balance, or lack thereof, between staff workload and patient needs, and the competence among both patients and staff. | 100% |
| 2020 | BMJ Open | Entezarjou et al. [14] | To explore staff experiences of working with a digital communication platform. | Flow Platform | Qualitative: Focus groups with GPs and nurses (n=19)  Primary healthcare centres (n=3) across Sweden, both rural and urban settings. | After a period of adjustment, it was seen as a useful communication tool by staff especially when combined with continuity of care. Staff acknowledged limitations regarding use by inappropriate patient populations, information overload and misinterpretation of text by both staff and patients. | 100% |
| 2020 | JMIR Medical Informatics | Entezarjou et al. [15] | To explore the interrater reliability between human physicians and an automated machine-learning (ML) based triage method. | Flow Platform | Quantitative: A naïve Bayes triage model created using data from digital medical histories, Sweden. Model was tested on 300 digital medical history reports and classification was compared with the majority vote of an expert panel of 5 primary care physicians (PCPs). | The model correctly classified 74% (138/186) of nonurgent cases, but only 42% (38/90) of urgent cases.  Interrater reliability in human triage using automated patient interviewing software reports is low (Cohen κ 0.20) and interrater triage reliability between a statistical model trained on automated patient interviewing software reports and a human panel was low (Cohen κ 0.17).  Limited possibility to use human decisions as a reference for ML to automate triage in primary care. | 80% |
| 2020 | JMIR Med Inform | Fernández et al. [16] | To analyse the sociodemographic factors that affect the likelihood of doctors using eConsulta | eConsulta | Quantitative: Retrospective cross-sectional analysis of administrative data (2451 doctors serving 220 centres, Catalan, Spain) | Doctors using eConsulta are 45-54 years of age, score higher than the 80th percentile on the quality of care index, have a high degree of accessibility, are involved in teaching, and work on a health team in a high socioeconomic urban setting. | 60% |
| 2020 | Findings from engagement report | Health Innovation Manchester [17] | To gather further feedback on The Greater Manchester (GM) Digital Primary Care Outcomes and Standards Framework | Digital Primary Health Care - unspecified | Mixed methods: Online survey (n=449 GM citizens), 4 focus group (n=26 citizens), interviews (n=5 citizens), Manchester, UK. | Digital tools supported access and enhanced patient experience, but there were barriers to use and safety concerns. | 0% |
| 2020 | Journal of Primary Care and Community Health | Johansson et al. [18] | To examine patients’ experiences with a new digital primary health care service using written dialogues. | Digital primary health care (DPHC) service using written dialogues – not specified | Quantitative: Survey: Patients (n=286), Region Skåne of southern Sweden. | Patients were satisfied, but expressed some uncertainty regarding the physician’s ability to assess correct care needs. | 60% |
| 2020 | Journal of Primary Care and Community Health | Johansson et al. [19] | To explore physicians’ experiences and satisfaction of digital primary health care. | Digital Doctor Reception (DDR) | Mixed methods: Survey and qualitative interviews with GPs in Sweden (n=6) | Using digital written patient dialogues complements traditional primary healthcare. GPs described satisfaction and having patients' self-described history was considered a significant patient safety factor. However, GPs felt that a predetermined symptom list were not suitable for all patients. | 60% |
| 2020 | Journal of the American Medical Informatics Association | Judson et al. [20] | To rapidly deploy a digital patient-facing self-triage and self-scheduling tool in a large academic health system to address the COVID-19 pandemic. | Patient portal-based COVID-19 self-triage and self-scheduling tool | Quantitative: San Francisco (UCSF) Health, US: large academic health system - 3 campuses, nearly 1000 inpatient beds, 9 primary care practices serving approx 90000 patients. | Tool was widely used by patients (completed 1129 times by 950 unique patients in the first 16 days). Of completed sessions, 315 (28%) were by asymptomatic patients, and 814 (72%) were by symptomatic patients. Data suggests unnecessary triage messages, phone calls, and in-person visits were prevented. | 40% |
| 2020 | International Journal of Medical Informatics | Kelley et al. [21] | To explore the effects of virtual visits on patient burden. | Online portal, virtual visits – not specified | Mixed methods: Evaluation of pilot implementation of virtual visits in Canada.  Patient interviews (n=17) and survey (n=427) | Virtual visits reduce work patients must do to manage their care by improving access, convenience, time needed for appointments and makes it easier to access info/support. Reduces perceived burden by improving continuity of care, experience and cost savings. | 40% |
| 2020 | Telemedicine and e-Health | Murray et al. [22] | To compare antibiotic prescribing, follow-up rates, and clinical outcomes between face-to-face visits at a retail clinic, nurse phone protocol encounters, and eVisits for the assessment and management of urinary symptoms and UTIs. | Symptom-specific questions via eVisit – not specified | Quantitative: Retrospective chart review of primary care empaneled patients at Mayo Clinic Rochester, US, of females, 18-65 years old, who sought care for urinary symptoms via phone, eVisit, or face-to-face.  450 encounters reviewed and compared for antibiotic prescribing rates, clinical outcomes, and 30-day follow-up rates. | Antibiotic prescribing rates for all three encounter types were similar. Referral for follow-up at initial encounter was more likely to be recommended from phone and eVisit encounters than face-to-face. No significant differences in follow-up rates or clinical outcomes noted between the three encounter types. | 80% |
| 2020 | Master Thesis, Lund University, Sweden | Peber and Wastfelt [23] | To develop and use an evaluation framework to study the impact of digi-physical healthcare on quality, efficiency, and patient and healthcare provider satisfaction. | Flow Platform | Mixed methods: Case study of primary care centres in Sweden (n=3), Staff survey (n=59) | Satisfaction is positively impacted by digi-physical care, while impact on quality and efficiency is uncertain. Ways of working influence what impact centres realise from digi-physical care. No impact on profits. | 20% |
| 2020 | Journal of Medical Internet Research | Segui et al. [24] | To annotate a random sample of teleconsultations from eConsulta, and to evaluate the level of agreement between healthcare professionals with respect to the annotation. | EConsulta | Quantitative: 20 GPs retrospectively annotated a random sample of 5382 cases managed by eConsulta (Catalonia, Spain) | GPs considered that 80% of teleconsultations helped avoid an in-person visit, and that 65% of the time, the patient would have made an in-person visit in the absence of eConsulta. Most frequent uses were for management of test results (27%) and management of repeat prescriptions (24%). Mixed degree of agreement among professionals. | 80% |
| 2020 | Journal of Medical Internet Research | Segui et al. [25] | To assess the ability of using eConsulta to reduce the number of face-to-face visits to primary care teams | EConsulta | Quantitative: 18 GPs classified 2268 cases managed with eConsulta and indicated whether the teleconsultation reduced the number of face-to-face visits.  (Catalonia, Spain) | GPs view eConsulta as having the potential to resolve patient queries for every type of consultation. Avoided need for a face-to-face visit in 87.9% of cases.  Ease of access increased the demand for health care support in 27.7% of cases. | 60% |
| 2020 | Health Informatics | Stamenova et al. [26] | To evaluate uptake of a platform for virtual visits in primary care, examine patient and physician preferences for virtual communication, and report on characteristics of visits and patients experiences | 2 platforms: Novari or Think Research | Quantitative: Retrospective cohort study. Primary care practices (n=326) in 5 regions of Ontario, Canada. 14,291 registered patients. | 44% of registered patients and 60% of registered providers used the platform at least once. Among patient users 51% completed at least one virtual visit. 81% of virtual visits required no follow-up and 99% of patients reported that they would use virtual care services again. | 40% |
| 2020 | BMJ Open | Zanaboni and Fagerlund [27] | To explore patient use and experiences with 4 digital health services, including e-consultation. | e-consultation integrated with patient record – not specified | Quantitative:  Online survey of portal users (n=2043), Norway. | High proportion of users are women, younger adults, digitally active and high education levels. For non-clinical inquiries, most respondents (60%) thought it easier to write electronic messages than communicate by phone. For clinical enquiries, patients agreed that e-consultation could lead to a better follow up (72%) and improved quality of treatment (58%). | 60% |
| 2019 | Transforming Government: People, Process and Policy | Andersen et al. [28] | To explore digital divide gaps and the impacts of online communication on the overall costs of health care. | E-visits: System not specified | Quantitative: Case study of online health care communication in Denmark. Analysis of e-visit population data between patients and GPs (3,500 e-visits) 2009-2015. | Increase in e-visits from 2009 to 2015: 1.3 million to 5.6 million (all ages). Higher use in women and ages 25-65 years.  Digital divide gaps exist in online healthcare communication, but uptake of e-visits does not widen socio-economic, gender or age gaps.  No associated decrease in overall number of consultations, no significant reduction of other consultations, no associated cost reductions. | 80% |
| 2019 | British Journal of General Practice | Eccles et al. [29] | To explore use of ‘AskMyGP’, and patients’ perspectives and experiences | AskMyGP Version 2 | Mixed methods: Retrospective analysis of ‘AskMyGP’ data from 5447 patients (9 UK practices) and free-text comments left by patients (n=569). | Highest levels of use were females (65.5%, n=3570) and ages 25–34. Common reasons for use: medication-related enquiries, administrative requests, and to report a specific symptom. Patients suggested advantages to using the platform (e.g. convenience), but these did not extend to all users. | 60% |
| 2019 | International Journal of Medical Informatics | Ekman et al. [30] | To provide a comprehensive review of the development of digital care in Sweden and how it compares with other types of primary care | Digital care platform – not specified | Quantitative: Descriptive analysis of national coverage data on the utilization of digital care by sex, age, place of residence, socioeconomic status, and most common diagnoses. | Digital primary care in Sweden has increased rapidly over the past two years (more than 30,000 digital consultations per month). Digital care differs to traditional care: users are generally younger and seek for different conditions. Digital care is similar to traditional care: utilisation is higher in metropolitan areas compared with rural areas. Similar to general health care use, there is a negative correlation between use of digital care and socioeconomic status. | 80% |
| 2019 | BMJ Open | Fagerlund et al. [31] | To explore GPs’ perceptions towards use of four digital health services, including e-consultation | e-consultation - not specified | Qualitative: Interviews with 9 GPs in Norway | Use of digital services in primary care in Norway is growing, although use of text-based e-consultations is limited. Most GPs were positive about all four services, but had scepticism regarding their effects. Advantages for GP offices included reduced phone load, increased efficiency, released time for medical assessments, less crowded waiting rooms and more precise communication. Benefits for patients were increased flexibility, autonomy and time and money savings. | 100% |
| 2019 | Mayo Clinic Proceedings | Hertzog et al. [32] | To compare diagnostic accuracy between primary care E-Visit and face-to-face encounters for low-acuity illnesses. | E-Visit program accessible through a patient's health portal - not specified | Quantitative: Cross-sectional retrospective analysis of electronic health records in a primary care setting in Texas, US (E-Visit n=490 and face-to-face n=2201). | Diagnostic accuracy for low-acuity illnesses in this population was equivalent between E-Visit and face-to-face encounters. | 40% |
| 2019 | Evaluation Report | IPSOS MORI [33] | What is the impact of Babylon GP at Hand on: registered patients; the wider health system; the workforce? | Babylon GP at Hand mobile and web app (BGPaH). | Mixed methods evaluation: Patient survey (n=1,452), case studies of practices (n=3), economic evaluation, qualitative interviews with staff (n=14) and patients (n=34) analysis of secondary data (North West London, UK. | Meets the needs of a specific segment of the population for a limited set of needs. Unclear whether the service is valid for a more diverse and multi-morbid population.  Attractive offer for some GPs, but unclear net impact on the wider workforce.  No evidence that BGPaH has an impact (positive or negative) on wider health service use. | 60% |
| 2019 | Online Consultations Research Report | NHS England and NHS Improvement [34] | To understand views and feelings of GP practice staff and the general public about online consultations. | Online consultations – not specified | Mixed methods, England, UK  Public: Online survey (n=3,066), street surveys in nine locations (n=1,674), 17 focus groups.  GPs and practice staff: Online survey (n=1,529), interviews with GPs (n=6) across the English regions | GPs significantly more likely to resist the idea of offering innovative new online services (57%) compared to any other job role.  Large, or group-sized practices (25,000+), likely to be more willing to offer new services compared to any other list size.  Patients aged 55-74 see lack of face-to-face contact as a barrier.  Low number of patients reporting experience of an online consultation (2.8% in survey, 1% in street survey). | 40% |
| 2019 | Journal of the American Board of Family Medicine | Peabody et al. [35] | To examine the prevalence of family physicians providing e-visits and associated factors. | e-visit - not specified | Quantitative: National cross-sectional practice questionnaire for practicing US family physicians (n=7580) | Fewer than 10% of family physicians provided e-visits. Reimbursement may be a barrier to providing e-visits. | 60% |
| 2018 | The British Journal of General Practice | Atherton et al. [36] | To understand how, under what conditions, for which patients and in what ways, alternatives to face-to-face consultations present benefits and challenges to patients and practitioners in general practice. | eConsult, AskMyGP | Qualitative: Ethnographic case studies in 8 UK general practices. Staff (n=45) and patients and carers (n=39) were interviewed | Implementation often not well thought through. Differing practice between team members. Patients reported benefits of convenience and access.  Staff and some patients felt the face-to-face consultation was the ideal. | 100% |
| 2018 | Health Services and Delivery Research (report) | Atherton et al. [37] | To understand why, how and with what consequences the use of telephone, e-mail or internet-based systems alongside face-to-face consultations in general practice. | eConsult, AskMyGP | Mixed methods: Multiple case study design  UK GP practices (n=8)  Staff (n=48) and patients and carers (n=39) were interviewed | Vast majority of consultations (80%) are still conducted face to face. E-consultations very rarely used (0.22% and 0.23% of consultations in practices that offer them). | 80% |
| 2018 | British Journal of General Practice | Banks et al. [38] | To evaluate whether an e-consultation system improves the ability of practice staff to manage workload and access. | eConsult | Qualitative: Interview study in general practices in the West of England UK that piloted an e-consultation system for 15 months (23 participants across 6 GP practices). | Routine e-consultations offered benefits for the practice as they could be completed without direct contact between GP and patient. However, most e-consultations resulted in a follow up telephone or face-to-face GP appointment due to insufficient information in the e-consultation. This was perceived as increasing workload and providing some patients with an alternative route into the appointment system. | 100% |
| 2018 | Management science | Bavafa et al. [39] | To estimate the impact of e-visit usage on visit frequency of office and phone encounters as well as on patient health outcomes. | Unspecified | Quantitative: Panel data set from a large healthcare system in the US (96,566 patients):  standard difference-in-differences analysis, matching on multiple dimensions and instrumental variable analysis. | e-visits trigger about 6% more office visits, with mixed results on phone visits and patient health. These additional visits come at the sacrifice of new patients: physicians accept 15% fewer new patients each month following e-visit adoption. | 40% |
| 2018 | BMJ Open | Carter et al. [40] | To evaluate the feasibility, acceptability, and effectiveness of WebGP | WebGP (now eConsult) | Mixed-methods: Evaluation of General practices in Northern, Eastern and Western Devon UK.  6 practices provided consultations data; 20 GPs completed case reports (regarding 61 e-consults); 81 patients completed questionnaires; 5 GPs and 5 administrators were interviewed. 81/231 patients completed a postal survey | WebGP uptake during the evaluation was small with no discernible impact on practice workload. GPs judged 41/61 (72%) WebGP requests to require a face-to-face or telephone consultation. While largely acceptable within practice, introducing e-consults had potential for adverse interactions with pre-existing practice systems. WebGP appeared broadly acceptable to patients regarding timeliness and quality/experience of care provided. | 60% |
| 2018 | International Journal of Environmental Research and Public Health | Cowie et al. [41] | To examine the impact of eConsult on patients, GP surgery, and staff; time implications; cost/effectiveness; barriers and facilitators to future implementation | eConsult  (Asynchronous) | Mixed methods: 11 GP practices across 4 Scottish NHS boards, UK.  Quantitative data: log data from eConsult use, patient survey data (completed by 6.5% of eConsult users).  Qualitative data: interviews (n=44) and 1 focus group (n=4) with GP staff, free text within patient survey (n=291). | Expectations that eConsult would offer an additional and alternative method of accessing GP services were largely met.  Lack of patient and staff engagement, insufficient support, and lack of protocols around processes were barriers. | 40% |
| 2018 | BMJ Open | Farr et al. [42] | To examine patient and staff views, experiences, and acceptability of an online consultation system and ask how the system and its implementation may be improved. | eConsult | Mixed-method evaluation, primary care practices in SW England, UK.  Qualitative interviews (23 practice staff in six practices).  Patient survey data for 756 e-consultations from 36 practices, with free-text survey comments from 512 patients.  Anonymised patients' records for 485 e-consultations from eight practices. | Most e-consultations resulted in either follow-on phone (32%) or face-to-face appointments (38%) and GPs felt that this duplicated their workload. Patient satisfaction of the system was high, a minority were dissatisfied with practice communication. | 60% |
| 2018 | Telemedicine and e-Health | Penza et al. [43] | To conduct a retrospective record review of patients who received care through an eVisit for a minor acute illness and to review 30-day outcomes of follow-up, emergency department visits, hospitalisations, and death for the same or related conditions. | Secure patient portal, e-visit – not specified | Quantitative: Retrospective record review (n=1009). Mayo clinic, US. Patients who had an e-visit within the study period were reviewed for 30 day outcomes of follow-up, ED visits, hospitalisations and death. | 1,009 eVisits analysed: 34% had follow-up within 30 days, with a follow-up rate of 20%.  Most e-visits for minor acute illnesses can be completed without any further interaction with the healthcare system. | 40% |
| 2018 | Health Affairs | Player et al. [44] | To evaluate an e-visit program in a US academic medical center setting. | Epic’s MyChart patient portal – e-visit | Quantitative: Retrospective chart review for nonemergent  acute care of adults in the period December 2015–July 2017 at the Medical University of South Carolina, US (1,565 e-visits) | Most patients were female (80.2%) and ages 18–44 (55.3%). Majority (81.5%) of in-person follow-ups did not result in diagnosis changes. More than 90% of the 665 patients surveyed after an e-visit reported a positive experience. Most patients (92%) reported that the e-visit had replaced an in-person visit. | 20% |
| 2017 | British Journal of General Practice | Casey et al. [45] | To explore the introduction of Tele-Doc and how it shapes working practices. | Tele-Doc | Mixed methods: Case study in a UK inner-city general practice.  Interviews with staff (n=7). | Low patient uptake. Consultation was redistributed to patients and administrators, sometimes causing misunderstandings. GPs welcomed varied modes of consulting, but the aspiration of improved efficiency was not realised. | 40% |
| 2017 | BMJ Open | Edwards et al. [46] | To describe: who used the system, when and why; and the National Health Service costs associated with its use. | eConsult | Quantitative: Evaluation of a pilot study. Primary care practices in South West England, UK: 36 General practices, 396 828 patients.  Data: routinely available data from Public Health England, website analytics data, random sample of patient data (users of e-consultations) from 8 practices. | 7472 patients completed an e-consultation. Women more likely to use than men (64.7% vs 35.3%), users had a median age of 39 years (IQR 30–50). Most common reason for an e-consultation was an administrative request (22.5%) followed by infections/ immunological issues (14.4%). Most common outcome was a face-to-face (38%) or telephone consultation (32%). Average cost of an e-consultation was £36.28, primarily triage time and resulting face-to-face/telephone consultations. | 80% |
| 2017 | Journal of Medical Internet Research | McGrail et al. [47] | To assess users and providers of virtual visits, including reasons patients give for use. To assess the influence of virtual visits on overall primary care use and costs | Virtual visits- not specified | Quantitative: Patient survey (n=399). Analysis of administrative health care data. British Columbia, Canada. | 7286 virtual visits, involving 5441 patients and 144 physicians. Younger patients and physicians more likely to use virtual visits, no differences by sex.  93.2% of patients said their virtual visit was of high quality; 91.2% reported it was “very” or “somewhat” helpful.  Virtual visits have the potential to decrease costs by approx Can $4 per quarter, but that benefit is most associated with seeing a known provider. | 60% |
| 2016 | London Journal of Primary Care | Lawless et al. [48] | To investigate different approaches to improving access to general practice and assess the impact on (i) patient experience, (ii) practice staff experience and (iii) activity in A&E and walk-in centres. | Online self-help advice and online consultation | Mixed methods: evaluation of 12 practices in Greenwich CCG, UK.  Pre (N=535) and post (N=1277) survey with practice users. Interviews with staff (N=21). | None of the approaches were overwhelmingly successful in improving patient experience of access or reducing practice workload.  Low uptake of online consultations, with decreasing use (therefore limited effect on access). | 60% |
| 2016 | Interim Update Report | Matheson [49] | Determine: patterns of use and outcomes of WebGP; impact on patient and staff experience; barriers and drivers to implementation; extent of embedding; and  cost effectiveness. | WebGP (now eConsult) | Mixed methods:  Evaluation of GP practices using WebGP (N=2), in Dorset and West Hampshire, UK. Data analysed from WebGP and e-consults usage and outcomes.  Survey of patients (N=?) and staff (N=8), focus groups with staff (N=7). | Patients positive about WebGP but made suggestions for improvement as staff responses were not always within one working day. Views of staff were polarised. Nearly 1/5 of e-consults were logged by patients aged >71. 28% e-consults were admin-only queries. | 20% |
| 2015 | MEDINFO 2015: eHealth-enabled Health (Conference paper) | Bertelsen and Petersen [50] | To report on selected findings from a Danish national survey of citizens’ perception and use of ICT for health care. Focus is on citizens’ use of ICT and communication with their GP. | E-consultation - not specified | Quantitative: National survey, questionnaire to a population sample (n=1058), Denmark | 65% of adult citizens or their relatives have used ICT to communicate with their GP (52% used e-consultation).  Data supports assumption that the higher the education people have, the more likely they are to use ICT for their health care. | 40% |
| 2015 | Evaluation Report | NHS England [51] | Focuses on three key national programme objectives, including to increase the range of contact modes. | WebGP (now eConsult) | Mixed methods: national evaluation of 20 pilot sites in England, UK, covering 1,100 general practices and 7.5 million patients (Six pilots introduced online patient diagnostic tools, including e-consultations).  Interviews with pilot leaders, partners and stakeholders, online survey with staff, monthly data on key services and innovation (n=?) | The pilot provided 470 on-line consultations. Mixed reception from GPs and patients. Bristol: 13 practices adopted e-consultations and the trial was seen as a success. In Brighton and Hove and Southwark some GPs were apprehensive about being inundated with requests. This led to reluctance to implement the system. Care UK implemented a diagnostic and e-consultation system at all eight of its practices but patients preferred telephone access. | 20% |
| 2014 | International Journal of Medical Informatics | Jung and Padman [52] | To learn characteristics of online healthcare consumers and understand their patterns of adoption and usage of online clinical consultation services. | eVisit (MyChart/Epic Corporation) | Quantitative: Analysis of the deployment of eVisits within a patient portal at four primary care clinics in Western Pennsylvania, US (2152 patients) | 324 patients used eVisits. On average, eVisit adopters are younger and predominantly female.  Younger, female patients have higher adoption hazard, but gender does not affect the decision of adopting early vs. late.  Patients with more complex health issues are more likely to use the service. | 80% |
| 2014 | Telemedicine and e-Health | North et al. [53] | To investigate the potential impact of portal messaging on a primary care practice. To examine a subgroup of high message utilising patients and determine whether increased portal communication is associated with fewer face-to-face visits. | Mayo Clinic Patient Portal, e-visit | Quantitative: Mayo Clinic, US. Retrospective cohort study. 2,357 primary care patients who used electronic messaging | No significant change in face-to-face visit frequency was observed following implementation of portal messaging. | 60% |
| 2013 | Health Affairs | Bishop et al. [54] | How can primary care practices use electronic communication to manage clinical issues traditionally managed during office visits?  What are the advantages and disadvantages for patients, physicians, and practices?  What are the barriers and facilitators to implementation? | Only one group used a formal e-visit program (not specified), others used non-structured input where patients entered text like an ordinary e-mail | Qualitative: Interviews over 4 months with 21 US medical groups and 6 leaders from national and regional health plans | Electronic communication was perceived to be a safe, effective, efficient means of communication that improves patient satisfaction and saves patients time, but that increases the volume of physician work unless office visits are reduced. In most cases the number of office visits did not decrease. | 80% |
| 2013 | JAMA Internal Medicine | Mehrotra et al. [55] | To compare care at e-visits and office visits for sinusitis and UTI. | e-visits through patient health record internet portal – not specified | Quantitative: Study of e-visits and office visits at 4 x primary care practices, US. | Of the 5165 visits for sinusitis, 465 (9%) were e-visits. Of the 2954 visits for UTI, 99 were e-visits (3%). Physicians less likely to order a UTI-relevant test at an e-visit (8% e-visits vs 51% office visits; P < .01)  No difference in how many patients had a follow-up visit for each condition.  Physicians more likely to prescribe an antibiotic at an e-visit for either condition.  During e-visits physicians less likely to order preventive care.  Data suggests e-visits could lower health care spending. | 40% |
| 2013 | Telemedicine Journal and a-Health | Mehrotra et al. [56] | To compare e-visit and office visit users for 2 conditions: sinusitis and UTI | eVisit (MyChart/Epic Corporation) | Quantitative: Electronic medical record analysis of demographic data.  5165 sinusitis visits (9% eVisit) and 2954 UTI visits (3% eVisit). Primary care practices (n=4), Pittsburgh US. | Variables most strongly associated with a patient initiating an eVisit versus an office visit: age, UTI, and longer travel distance to clinic. Higher income was not associated with higher eVisit use. | 40% |
| 2013 | Journal of American Medical Informatics Association | North et al. [57] | To examine how often patients are using secure messaging and e-visits for acute, high risk symptoms. | Mayo Clinic Patient Portal, e-visit | Quantitative: Retrospective content analysis of 7322 messages (secure messages and e-visits), Mayo clinic, US.  Looking for deaths within 30 days and hospitalisations/ED visits within 7 days. Analysis for high risk symptoms. | Patients use portal messages 3.5% of the time for potentially high-risk symptoms. Death, hospitalisation or an ED visit was an infrequent outcome following secure message/e-visit. Screening subject line for high-risk symptoms was not successful in identifying high risk message content. | 60% |
| 2011 | Telemedicine and e-Health | Albert et al. [58] | To investigate outcomes associated with e-Visits: whether patients receiving diagnoses receive appropriate care or need to return to the doctor. | eVisit (MyChart/Epic Corporation) | Quantitative: Survey of the first 156 e-Visit users (n=121) from a large family medicine practice (Pittsburgh, US). Medical records for patients making e-Visits were reviewed to examine need for follow-up care within 7 days. | Majority of patients (57%) were diagnosed without need for follow-up beyond a prescription; 75% of patients thought the e-Visit was as good or better than an in-person visit, 11.6% felt their concerns were incompletely addressed. In a review of medical records, 16.9% had a follow-up visit within 7 days, mostly for the same condition. | 80% |
| 2011 | AMIA Annual Symposium Proceedings (conference paper) | Jung et al. [59] | To analyse the key features that distinguish early adopters of eVisits from portal consumers | eVisit (MyChart/Epic Corporation) | Quantitative: Usage data of an online health portal in a healthcare provider in Western Pennsylvania, US (four ambulatory practices). | Out of 10,532 portal users, the 336 patients who submitted 446 eVisits were younger, predominantly female, in poorer health condition.  Practice indicator is a significant predictor of eVisit usage. Insurance coverage for e-visits significantly contributes to increased usage. | 60% |
| 2010 | Mayo Clinic Proceedings | Adamson and Bachman [60] | To study the use of e-visits in a primary care setting | Intuit | Quantitative: Pilot study, US. 24 months duration.  4x clinics, 56 clinicians. Data were collected from e-visits | 4282 patients registered.  2531 e-visits.  E-visits primarily by women during working hours. Range of conditions (294) were dealt with. 411 (16%) encounters previously unbillable now billable. 1012 (40%) did not need clinic visits. 324 (13%) led to scheduled in-person appointments. | 20% |
| 2010 | 2^nd^ international Conference on eHealth, Telemedicine and Social Medicine | Nijland et al. [61] | To assess whether an e-consultation service is beneficial and efficient for giving support on health related requests. | MedicInfo | Quantitative: Evaluation of e-consultation in the Netherlands.  Content analysis of requests (n=222) | E-consultation used for health requests on non-urgent, minor ailments. Also used for decision support on whether a visit to a doctor was necessary.  Users predominantly women (68%), mean age 44yrs. 8% submitted for others e.g. child/partner. | 60% |
| 2010 | Studies in Health Technology and Informatics | Padman et al. [62] | To examine the adoption and use of a pilot eVisit project | eVisit (MyChart/Epic Corporation) | Mixed methods: Pilot study evaluation. One primary care clinic (3 locations), US. 7 health conditions.  Data analysed from: eVisits (n=152), messages (n=417), patient satisfaction surveys (n=28) and physician satisfaction surveys and interviews (n=11) | 126 patients and 11 physicians used the service.  4% of new portal users using e-visits increased to 25% through study period. 50% were ages 36-55. Women more frequent users. Few exchanges were needed to resolve a request. | 60% |
| 2010 | Population Health Management | Rohrer et al. [63] | To compare the odds of being a cost outlier during a 6-month period after either an online visit or an in-person visit. | Secure web-based portal, online visit – not specified | Quantitative: Comparison of patients who had an online visit (n=390) vs regular office care (n=376), US  Outliers defined as patients for whom standard costs exceeded the 75^th^ percentile during a 6-month period after the initial visit. | 21.2% of online visitors were cost outliers (versus 28.5% in the standard visit group).  Online visits appeared to reduce medical costs for patients during a 6-month period after the visit. | 60% |

1. See Appendix 5 for further details
