## Appendix 5 for "Understanding how the design and implementation of Online Consultations influence primary care outcomes: Systematic review of evidence with recommendations for designers, providers, and researchers"

**Appendix 5: Description of Online Consultation systems studied**

| **Software program (if stated)** | **Description** | **Relevant features from Table 1** | **Papers** |
| --- | --- | --- | --- |
| eConsult (previously WebGP) | Online platform which includes symptom checker, pharmacy advice, administrative service, and e-consultation. To submit an e-consultation, patients complete an online form to provide a structured medical account of their condition. Once received, clinicians contact the patient, usually by telephone. | - One-way communication only - Multiple choice questionnaires with optional free-text - No integration - No artificial intelligence | [1-9] |
| Tele-Doc | A condition-specific online questionnaire with five sections, all of which must be completed. Received by the practice as an email attachment and dealt with according to locally specified organisational routines. | - One-way communication only - Multiple choice questionnaires with optional free-text - No integration - No artificial intelligence | [10] |
| eVisit or UPMC HealthTrak (MyChart/Epic Corporation) | Patient selects a condition, completes the structured documentation and submits the eVisit via the portal. Support staff review and forward the submission to a physician. | - Asynchronous two-way communication - Multiple choice questionnaires with optional free-text - Electronic health record integration - Adapting patient questions during query submission - Pre-programmed logic | [11-15] |
| Flow Platform (Doctrin AB) | Patients choose among a prespecified list of queries and access a structured patient interviewing software, where patients can briefly formulate ideas, concerns, and expectations, and subsequently answer a symptom-specific multiple-choice survey. The software selects suitable subsequent survey questions based on the patient’s answers. A healthcare provider (usually a nurse) then contacts the patient. | - Asynchronous two-way communication - Multiple choice questionnaires with optional free-text - No integration - Adapting patient questions during query submission - Artificial intelligence method: unclear | [16-19] |
| Mayo Clinic Patient Portal | Computer directed interview resulting in a structured message to the provider with information pertinent to the new symptom. Providers respond to e-visits by text (secure message) or telephone. | - Asynchronous two-way communication - Multiple choice questionnaires only - No integration - No artificial intelligence | [20, 21] |
| Intuit | Patients enter reported problem (e.g. back pain) and then answer questions one at a time. Questions branch such that history is organized into a readable clinical format. Including validated diagnostic instruments, photo uploads, and additional free-text following completion of questionnaire. | - Asynchronous two-way communication - Multiple choice questionnaires with optional free-text - No integration - Adapting patient questions during query submission - Pre-programmed logic | [22] |
| Patient portal self-triage and self-scheduling tool for COVID-19 | Automated symptom assessment, triage, and appointment scheduling for patients with concerning symptoms or questions about COVID-19. After answering a series of questions, patients are segmented into risk categories and directed to 1 of 4 endpoints. Basic demographic information is automatically populated from the patient’s medical record. Clinicians only review encounters if patients call the triage hotline or clinic or make an appointment. | - One-way communication only - Multiple choice questionnaires only - Electronic health record integration - Appointment scheduling - Adapting patient questions during query submission - Prioritising patient queries based on clinical urgency - Signposting patients to the most appropriate care provider - Pre-programmed logic | [23] |
| Babylon GP at Hand | Includes an ‘AI’ triage system based on “chat-bot” symptom-checker. Patients are provided with advice, which might be self-care, message a clinician or have a telephone/video call. | - Asynchronous two-way communication - Synchronous real-time communication - Multiple choice questionnaires with optional free-text - Electronic health record integration - Prioritising patient queries based on clinical urgency - Signposting patients to the most appropriate care provide - Artificial intelligence method: unclear | [24] |
| Dr iQ | Algorithms with screening questions. Submissions are reviewed by a group of clinicians. | - Asynchronous two-way communication - Synchronous real-time communication - Multiple choice questionnaires only - No integration - Adapting patient questions during query submission - Pre-programmed logic | [25] |
| askmyGP | An online form detailing background and query, which is reviewed by administrative and clinician staff. | - One-way communication only - Multiple choice questionnaires with optional free-text - No integration - No artificial intelligence | [1, 9, 26] |
| MedicInfo | Free-text request sent to the GP. When answering, clinicians use one of four possible protocols. | - Asynchronous two-way communication - Unstructured free-text only - No integration - No artificial intelligence | [27] |
| eConsulta | Users choose which health professional to direct their enquiry to and attach files, while keeping a record of previous interactions. | - Asynchronous two-way communication - Unstructured free-text only - Electronic health record integration - No artificial intelligence | [28-30] |
| Digital Doctor Reception (DDR) | Patients complete a “medical history,” including cause of contact, background disease/s, and current inconvenience. Record form is handled digitally by a GP and continued through written digital dialogue. | - Asynchronous two-way communication - Multiple choice questionnaires with optional free-text - Electronic health record integration - No artificial intelligence | [31] |
| Think Research platform and Novari platform | Patients can request visits to be completed through secure messaging, audio and/or video. Once a patient request is sent, the primary care provider receives a notification on the platform and through email. They log into the platform to see the request and respond through any of the three modalities. | - Asynchronous two-way communication - Synchronous real-time communication - Unstructured free-text only - No integration - No artificial intelligence | [32] |
| Epic’s MyChart patient portal | Patient chooses the e-visit condition that matches their symptoms and fills out a condition-specific questionnaire. Once the questionnaire is complete, the provider reviews it and responds to the patient with a treatment plan. | - Asynchronous two-way communication - Multiple choice questionnaires only - Electronic health record integration - No artificial intelligence | [33] |
| Docly online consultation | Patients select their health problem, then fill in a guided questionnaire about their symptoms. Decision support algorithms recommend the most appropriate outcome to the clinician based on questionnaire answers, before either providing immediate advice or a further assessment is made by a GP. Health problems requiring more complex care are referred to specialist care services. Emergency situations are flagged by the smart algorithms, with patients receiving advice to contact urgent care services. | - Asynchronous two-way communication - Multiple choice questionnaires only - No integration - Prioritising patient queries based on clinical urgency - Signposting patients to the most appropriate care provider - Pre-programmed logic | [34] |
| Unnamed (chat automation system) | Patients specify which medical problem they are seeking advice for and answer predefined questions derived for this problem, and then describe their symptoms using a chat with nurses and/or physicians. | - Asynchronous two-way communication - Multiple choice questionnaires with optional free-text - No integration - Adapting patient questions during query submission - Artificial intelligence method: unclear | [35] |
| Unnamed (eVisit) | Patient submits answers to predetermined questions, which are reviewed by Advanced Practice Providers. | - Asynchronous two-way communication - Multiple choice questionnaires only - No integration - Adapting patient questions during query submission - Pre-programmed logic | [36] |
| Unnamed (eVisit) | Patient selects their symptom/ healthcare concern from a menu and is directed to a symptom-specific structured set of algorithmic questions. Nurses review submissions. | - Asynchronous two-way communication - Multiple choice questionnaires only - Electronic health record integration - Adapting patient questions during query submission - Pre-programmed logic | [37] |
| Unnamed (Online visits) | Using a branching logic program, medical history is obtained, along with patient-provided medication list, pharmacy of choice, and vital signs. Once the specific request is completed by the patient, it is sent to the patient’s physician for review. | - Asynchronous two-way communication - Multiple choice questionnaires only - No integration - Adapting patient questions during query submission - Automated history taking - Pre-programmed logic | [38] |
| Unnamed (eVisits) | Patient-completed symptom specific question sets sent to a pool of Advanced Practice Providers via a secure online patient portal. | - Asynchronous two-way communication - Multiple choice questionnaires only - No integration - No artificial intelligence | [39] |
| Unnamed (e-consultation) | Patients initiate an e-consultation by logging into a level 4 security portal, where a written message can be sent to the GP. | - Asynchronous two-way communication - Unstructured free-text only - No integration - No artificial intelligence | [40] |
| Unnamed (Digital primary health care (DPHC) service) | Patients enter medical history, reason for contact, background disease(s), and current inconvenience. Record form is then digitally reviewed by a general practitioner (GP), who contacts the patient. | - Asynchronous two-way communication - Multiple choice questionnaires with optional free-text - No integration - No artificial intelligence | [41] |
| Unnamed (chat-based and automated medical history–taking service) | Before the chat, an automated medical history–taking service is offered, where the patient responds to a questions about their chief complaint and current health status. | - Asynchronous two-way communication - Multiple choice questionnaires with optional free-text - No integration - Adapting patient questions during query submission - Pre-programmed logic | [42] |
| Unnamed (e-visits) | Patients log into their secure personal health record internet portal and answer a series of questions about their condition. This written information is sent to the physicians, who make a diagnosis, order necessary care, put a note in the patients' electronic medical records, and reply to the patients via the secure portal within several hours. | - Asynchronous two-way communication - Patient query format: unclear - No integration - No artificial intelligence | [43] |
| Unnamed (e-consultation) | An online text-based clinical consultation with the GP, used only for known health conditions. Accessed by secure login to the national health portal. Includes a medical assessment of the patient’s request and is considered complete when the doctor has processed the inquiry and provided an answer. | - Asynchronous two-way communication - Patient query format: unclear - Electronic health record integration - No artificial intelligence | [44] |
| Unnamed (virtual visits) | Patients log in to an online portal to initiate a visit. Their primary care provider accepts the visit and responds in the most appropriate modality. The platform is primarily intended for two-way communication between the patient and the provider, however, patients can also request copies of lab results and prescription renewals. | - Asynchronous two-way communication - Synchronous real-time communication - Patient query format: unclear - No integration - No artificial intelligence | [45] |
| Unnamed (online consultations) | Allows patients to remotely and asynchronously contact a GP using a computer, smartphone, or tablet to ask questions and describe symptoms in writing. | - Communication mode: Unclear - Multiple choice questionnaires with optional free-text - No integration - No artificial intelligence | [46] |
| Unnamed (online consultations) | Allows patients to remotely and asynchronously contact a GP using a computer, smartphone, or tablet to ask questions and describe symptoms in writing. | - Communication mode: Unclear - Multiple choice questionnaires - No integration - No artificial intelligence | [46] |
| Unnamed (online consultations) | Allows patients to remotely and asynchronously contact a GP using a computer, smartphone, or tablet to ask questions and describe symptoms in writing. | - Communication mode: Unclear - Multiple choice questionnaires - No integration - Prioritising patient queries based on clinical urgency - Pre-programmed logic | [46] |
| Unnamed | Detailed description of features unavailable |  | [47-62] |
