## Appendix 6 for "Understanding how the design and implementation of Online Consultations influence primary care outcomes: Systematic review of evidence with recommendations for designers, providers, and researchers"

**Appendix 6: Quality appraisal of included studies using the Mixed Methods Appraisal Tool [1]**

| **Studies** |  | | **Question (see below for details)** | | | | | | | | | | | | | | | | | | | | | | | | |  |
| --- | --- | --- | --- | --- | --- | --- | --- | --- | --- | --- | --- | --- | --- | --- | --- | --- | --- | --- | --- | --- | --- | --- | --- | --- | --- | --- | --- | --- |
|  | **S1** | **S2** | **1.1** | **1.2** | **1.3** | **1.4** | **1.5** | **2.1** | **2.2** | **2.3** | **2.4** | **2.5** | **3.1** | **3.2** | **3.3** | **3.4** | **3.5** | **4.1** | **4.2** | **4.3** | **4.4** | **4.5** | **5.1** | **5.2** | **5.3** | **5.4** | **5.5** | **Overall score^[1]^** |
| ADAMSON ET AL. [2] | Y | Y |  |  |  |  |  |  |  |  |  |  |  |  |  |  |  | ? | N | ? | Y | N |  |  |  |  |  | 1*/20% |
| ALBERT ET AL. [3] | Y | Y |  |  |  |  |  |  |  |  |  |  |  |  |  |  |  | Y | N | Y | Y | Y |  |  |  |  |  | 4****/80% |
| ANDERSEN ET AL. [4] | Y | Y |  |  |  |  |  |  |  |  |  |  |  |  |  |  |  | Y | Y | Y | Y | ? |  |  |  |  |  | 4****/80% |
| ATHERTON ET AL. [5] | Y | Y | Y | Y | Y | Y | Y |  |  |  |  |  |  |  |  |  |  |  |  |  |  |  |  |  |  |  |  | 5*****/100% |
| ATHERTON ET AL. [6] | Y | Y | Y | Y | Y | Y | Y |  |  |  |  |  |  |  |  |  |  | Y | N | Y | Y | Y | Y | Y | Y | Y | Y | 4****/80% |
| BANKS ET AL. [7] | Y | Y | Y | Y | Y | Y | Y |  |  |  |  |  |  |  |  |  |  |  |  |  |  |  |  |  |  |  |  | 5*****/100% |
| BAVAFA ET AL. [8] | Y | Y |  |  |  |  |  |  |  |  |  |  | Y | Y | N | N | ? |  |  |  |  |  |  |  |  |  |  | 2**/40% |
| BERTELSEN AND PETERSEN [9] | N | ? |  |  |  |  |  |  |  |  |  |  |  |  |  |  |  | N | Y | Y | N | N |  |  |  |  |  | 2**/40% |
| BISHOP ET AL. [10] | Y | Y | Y | N | Y | Y | Y |  |  |  |  |  |  |  |  |  |  |  |  |  |  |  |  |  |  |  |  | 4****/80% |
| CAJANDER ET AL. [11] | Y | Y | N | N | Y | Y | Y |  |  |  |  |  |  |  |  |  |  |  |  |  |  |  |  |  |  |  |  | 3***/60% |
| CARTER ET AL. [12] | Y | Y | Y | Y | Y | Y | Y |  |  |  |  |  |  |  |  |  |  | Y | N | Y | N | Y | Y | N | Y | Y | N | 3***/60% |
| CASEY ET AL. [13] | N | ? | N | N | Y | Y | N |  |  |  |  |  |  |  |  |  |  |  |  |  |  |  |  |  |  |  |  | 2**/40% |
| COWIE ET AL. [14] | Y | N | Y | N | Y | Y | Y |  |  |  |  |  |  |  |  |  |  | Y | N | N | N | Y | Y | Y | Y | Y | N | 2**/40% |
| ECCLES ET AL. [15] | Y | Y | N | N | Y | Y | Y |  |  |  |  |  |  |  |  |  |  | Y | Y | Y | N | Y | Y | Y | Y | Y | Y | 3***/60% |
| EDWARDS ET AL. [16] | Y | Y |  |  |  |  |  |  |  |  |  |  |  |  |  |  |  | Y | N | Y | Y | Y |  |  |  |  |  | 4****/80% |
| EKMAN ET AL. [17] | Y | Y |  |  |  |  |  |  |  |  |  |  |  |  |  |  |  | Y | Y | Y | Y | ? |  |  |  |  |  | 4****/80% |
| ELDH ET AL. [18] | Y | Y | Y | Y | Y | Y | Y |  |  |  |  |  |  |  |  |  |  |  |  |  |  |  |  |  |  |  |  | 5*****/100% |
| ENTEZARJOU ET AL. [19] | Y | Y | Y | Y | Y | Y | Y |  |  |  |  |  |  |  |  |  |  |  |  |  |  |  |  |  |  |  |  | 5*****/100% |
| ENTEZARJOU ET AL. [20] | Y | Y |  |  |  |  |  |  |  |  |  |  | N | Y | Y | Y | Y |  |  |  |  |  |  |  |  |  |  | 4****/80% |
| FAGERLUND ET AL. [21] | Y | Y | Y | Y | Y | Y | Y |  |  |  |  |  |  |  |  |  |  |  |  |  |  |  |  |  |  |  |  | 5*****/100% |
| FARR ET AL. [22] | Y | Y | Y | Y | Y | Y | Y |  |  |  |  |  |  |  |  |  |  | Y | N | Y | N | Y | N | Y | Y | Y | N | 3***/60% |
| FERNANDEZ ET AL. [23] | Y | Y |  |  |  |  |  |  |  |  |  |  | Y | Y | Y | N | ? |  |  |  |  |  |  |  |  |  |  | 3***/60% |
| HEALTH INNOVATION MANCHESTER [24] | N | ? | ? | N | Y | N | N |  |  |  |  |  |  |  |  |  |  |  |  |  |  |  |  |  |  |  |  | 1*/20% |
| HEALTH INNOVATION MANCHESTER [25] | N | ? | ? | ? | N | N | N |  |  |  |  |  |  |  |  |  |  | ? | Y | ? | ? | ? | N | ? | Y | N | ? | 0*/0% |
| HERTZOG ET AL. [26] | Y | Y |  |  |  |  |  |  |  |  |  |  | N | Y | Y | N | ? |  |  |  |  |  |  |  |  |  |  | 2**/40% |
| IPSOS MORI [27] | Y | Y | Y | Y | N | Y | Y |  |  |  |  |  |  |  |  |  |  | Y | Y | N | N | Y | N | Y | Y | Y | ? | 3***/60% |
| JOHANSSON ET AL. [28] | Y | Y |  |  |  |  |  |  |  |  |  |  |  |  |  |  |  | Y | N | Y | N | Y |  |  |  |  |  | 3***/60% |
| JOHANSSON ET AL. [29] | Y | Y | Y | Y | Y | Y | Y |  |  |  |  |  |  |  |  |  |  | N | N | Y | Y | Y | Y | Y | Y | Y | N | 3***/60% |
| JUDSON ET AL. [30] | N | ? |  |  |  |  |  |  |  |  |  |  |  |  |  |  |  | ? | ? | Y | Y | N |  |  |  |  |  | 2**/40% |
| JUNG AND PADMAN [31] | Y | Y |  |  |  |  |  |  |  |  |  |  | Y | Y | Y | Y | ? |  |  |  |  |  |  |  |  |  |  | 4****/80% |
| JUNG ET AL. [32] | N | ? |  |  |  |  |  |  |  |  |  |  | N | Y | Y | Y | ? |  |  |  |  |  |  |  |  |  |  | 3***/60% |
| KELLEY ET AL. [33] | Y | Y | Y | Y | Y | Y | Y |  |  |  |  |  |  |  |  |  |  | N | N | Y | N | Y | Y | Y | Y | Y | N | 2**/40% |
| LANDGREN AND CAJANDER [34] | Y | N | Y | N | Y | Y | N |  |  |  |  |  |  |  |  |  |  |  |  |  |  |  |  |  |  |  |  | 3***/60% |
| LAWLESS ET AL. [35] | Y | Y | Y | Y | ? | Y | ? |  |  |  |  |  |  |  |  |  |  | Y | N | Y | Y | Y | Y | Y | Y | N | Y | 3***/60% |
| LEUNG AND QURESHI [36] | Y | Y | Y | Y | N | Y | Y |  |  |  |  |  |  |  |  |  |  | ? | N | Y | Y | ? | Y | Y | Y | Y | Y | 2**/40% |
| MATHESON [37] | Y | N | Y | Y | Y | Y | Y |  |  |  |  |  |  |  |  |  |  | ? | N | N | Y | N | Y | Y | Y | Y | N | 1*/20% |
| MCGRAIL ET AL. [38] | Y | Y |  |  |  |  |  |  |  |  |  |  | Y | Y | Y | N | ? | Y | Y | Y | N | Y |  |  |  |  |  | 3***/60% |
| MEHROTRA ET AL. [39] | Y | Y |  |  |  |  |  |  |  |  |  |  | N | Y | Y | N | ? |  |  |  |  |  |  |  |  |  |  | 2**/40% |
| MEHROTRA ET AL. [40] | Y | Y |  |  |  |  |  |  |  |  |  |  |  |  |  |  |  | N | N | N | Y | Y |  |  |  |  |  | 2**/40% |
| MURPHY ET AL. [41] | Y | Y | Y | Y | Y | Y | Y |  |  |  |  |  |  |  |  |  |  | Y | N | Y | Y | Y | N | N | N | Y | N | 1*/20% |
| MURRAY ET AL. [42] | Y | Y |  |  |  |  |  |  |  |  |  |  |  |  |  |  |  | Y | N | Y | Y | Y |  |  |  |  |  | 4****/80% |
| NIJHOF ET AL. [43] | Y | N |  |  |  |  |  |  |  |  |  |  |  |  |  |  |  | Y | Y | N | Y | N |  |  |  |  |  | 3***/60% |
| NIJLAND ET AL. [44] | Y | Y |  |  |  |  |  |  |  |  |  |  |  |  |  |  |  | Y | ? | Y | ? | Y |  |  |  |  |  | 3***/60% |
| NILSSON ET AL. [45] | Y | Y | Y | Y | Y | Y | Y |  |  |  |  |  |  |  |  |  |  |  |  |  |  |  |  |  |  |  |  | 5*****/100% |
| NHS ENGLAND [46] | N | ? |  |  |  |  |  |  |  |  |  |  |  |  |  |  |  |  |  |  |  |  | Y | ? | ? | ? | ? | 1*/20% |
| NHS ENGLAND [47] | N | ? | ? | ? | Y | Y | ? |  |  |  |  |  |  |  |  |  |  | ? | ? | Y | ? | Y | ? | ? | Y | Y | ? | 2**/40% |
| NORTH ET AL. [48] | Y | Y |  |  |  |  |  |  |  |  |  |  | N | Y | Y | Y | ? |  |  |  |  |  |  |  |  |  |  | 3***/60% |
| NORTH ET AL. [49] | N | ? |  |  |  |  |  |  |  |  |  |  | N | Y | Y | Y | ? |  |  |  |  |  |  |  |  |  |  | 3***/60% |
| PADMAN ET AL. [50] | Y | Y | Y | ? | Y | Y | ? |  |  |  |  |  |  |  |  |  |  | Y | Y | Y | Y | Y | Y | Y | Y | Y | N | 3***/60% |
| PEABODY ET AL. [51] | Y | Y |  |  |  |  |  |  |  |  |  |  | Y | Y | Y | N | ? |  |  |  |  |  |  |  |  |  |  | 3***/60% |
| PEBER AND WÄSTFELT [52] | Y | Y |  |  |  |  |  |  |  |  |  |  |  |  |  |  |  | N | N | N | Y | N |  |  |  |  |  | 1*/20% |
| PLAYER ET AL. [53] | N | ? |  |  |  |  |  |  |  |  |  |  |  |  |  |  |  | ? | N | ? | Y | ? |  |  |  |  |  | 1*/20% |
| PENZA ET AL. [54] | Y | Y |  |  |  |  |  |  |  |  |  |  | Y | Y | Y | ? | ? |  |  |  |  |  |  |  |  |  |  | 3***/60% |
| PENZA ET AL. [55] | N | ? |  |  |  |  |  |  |  |  |  |  | N | Y | Y | N | ? |  |  |  |  |  |  |  |  |  |  | 2**/40% |
| ROHRER ET AL. [56] | Y | Y |  |  |  |  |  |  |  |  |  |  | N | Y | Y | Y | ? |  |  |  |  |  |  |  |  |  |  | 3***/60% |
| SEGUI ET AL. [57] | Y | Y |  |  |  |  |  |  |  |  |  |  |  |  |  |  |  | Y | N | Y | Y | Y |  |  |  |  |  | 4****/80% |
| SEGUI ET AL. [58] | Y | Y |  |  |  |  |  |  |  |  |  |  | N | Y | Y | N | Y | Y | N | Y | Y | Y |  |  |  |  |  | 3***/60% |
| STAMENOVA ET AL. [59] | Y | Y |  |  |  |  |  |  |  |  |  |  |  |  |  |  |  | N | Y | Y | N | ? |  |  |  |  |  | 2**/40% |
| TARN ET AL. [60] | Y | Y |  |  |  |  |  |  |  |  |  |  | N | Y | Y | Y | ? |  |  |  |  |  |  |  |  |  |  | 3***/60% |
| TURNER ET AL. [61] | Y | N | Y | N | Y | Y | N |  |  |  |  |  |  |  |  |  |  |  |  |  |  |  |  |  |  |  |  | 3***/60% |
| WILSON ET AL. [62] | Y | Y | Y | N | Y | Y | Y |  |  |  |  |  |  |  |  |  |  |  |  |  |  |  |  |  |  |  |  | 4****/80% |
| ZANABONI AND FAGERLUND [63] | Y | Y |  |  |  |  |  |  |  |  |  |  |  |  |  |  |  | Y | N | Y | ? | Y |  |  |  |  |  | 3***/60% |

5.5. Do the different components of the study adhere to the quality criteria of each tradition of the methods involved?

*Each section is marked out of 100 (20 marks for each question). For mixed methods studies, the overall quality score is the lowest score of the study components.*

24.	Health Innovation Manchester. GM Digital First Primary Care: Patient and public insights: Workshop results. Report. Manchester: Health Innovation Manchester, 2021 Sept 2021. Report No.: 1 Contract No.: 1 Sept.

1. Colour coding for MMAT to distinguish low to high ratings: Red (20%), grey (40%), yellow (60%), blue (80%), green (100%)
