## Appendix 7 for "Understanding how the design and implementation of Online Consultations influence primary care outcomes: Systematic review of evidence with recommendations for designers, providers, and researchers"

**Appendix 7: Low confidence findings for objective 1**

| **Theme** | **Subtheme** |
| --- | --- |
| **Effective** (providing care based on scientific knowledge to produce better clinical outcomes) | **Neutral and increased antibiotic prescribing rates**  **Description:** The same or higher rate of prescribed antibiotics than through traditional consultations  **CERQual rating:** Low  **CERQual explanation:** Low adequacy and low coherence  **References:** [1-3], n=3  **Exemplar data:** Physicians were more likely to prescribe an antibiotic at an e-visit for either condition [sinusitis and UTI] [1] |
|  | **Reduced antibiotic prescribing rates**  **Description:** Fewer antibiotics prescribed through OCs  **CERQual rating:** Low  **CERQual explanation:** Low adequacy and low coherence  **References:** [4-6], n=3  **Exemplar data:** Patients evaluated F2F were more likely to be given an antibiotic prescription (72% [108/ 150]) when compared with those evaluated through e-visit (56% [84/150]; p = 0.004) [5] |
