## Appendix 8 for "Understanding how the design and implementation of Online Consultations influence primary care outcomes: Systematic review of evidence with recommendations for designers, providers, and researchers"

**Appendix 7: Low confidence findings for objective 2**

| **Theme** | **OC design feature or implementation** | **Outcome (from Table 2)** | **CERQual rating, references, and exemplar data** |
| --- | --- | --- | --- |
| **Condition**  (illness OC is used for) | **Sensitive issues** | **Patient-centeredness:**  Increased patient satisfaction (qualitative and quantitative) | **CERQual rating:** Low  **CERQual explanation:** Low adequacy  **References:** [1, 2], n=2  **Exemplar data:** Practices also perceived other patient benefits around the ability that patients have to articulate their concerns with less  fear of embarrassment:  ‘The feedback from patients I found really positive. I think they’ve really, you know, found it; I think they like it and a couple of them crystallise it around saying that it was a bit of an embarrassing problem and this almost allowed me to sort of hide behind. I haven’t got to have a whites of the eyes conversation with my GP. I can put  it in an e-mail and it feels very detached when I send it off and then I get the answer or result back without having to sort of embarrass myself so that’s worked well.’ (AM10). [2] |
|  | **Decreased complexity** | **Patient-centeredness:**  Increased patient satisfaction (qualitative and quantitative) | **CERQual rating:** Low  **CERQual explanation**: Low adequacy  **References:** [3, 4], n=2  **Exemplar data:** Patients in this e-Visit group clearly appreciated not having to travel to physician offices or trying to reach the doctor’s office by telephone to discuss what they considered routine symptoms only requiring a prescription. [4] |
|  | **Increased complexity** | **Adoption:** High adoption (qualitative and quantitative) | **CERQual rating:** Low  **CERQual explanation:** Low adequacy  **References:** [5, 6], n=2  **Exemplar data:** The more complex a patient’s medical background, the more likely they were to use the OC. [6] |
|  |  | **Adoption:** Low adoption (qualitative and quantitative) | **CERQual rating:** Low  **CERQual explanation:** Low adequacy and low coherence  **References:** [7], n=1  **Exemplar data:** The evaluation of Babylon GP at Hand demonstrated that the service is not being used by people with more complex health needs. [7] |
| **Technology**  (material properties of OC) | **Technical difficulties** | **Patient-centeredness:**  Decreased patient satisfaction (qualitative) | **CERQual rating:** Low  **CERQual explanation:** Low adequacy  **References:** [1, 8, 9], n=3  **Exemplar data:** Challenges with the OC, such as “glitches” or being “hard to navigate and fraught with errors” contributed to a negative experience for some patients. [1] |
|  | **Free text** | **Patient-centeredness:**  Decreased patient satisfaction (qualitative) | **CERQual rating:** Low  **CERQual explanation:** Low adequacy  **References:** [10], n=1  **Exemplar data:** When a free-text option was available, some patients struggled with how best to explain their issue owing to uncertainty about who they were writing to and who would read their enquiry. [10] |
|  | **One-way communication** | **Efficiency:** Increased workload (qualitative and quantitative) | **CERQual rating:** Low  **CERQual explanation:** Low adequacy  **References:** [11, 12], n=2  **Exemplar data:** Several interviewees suggested improvements to the webGP programme, such as a way of alerting patients to expect a call from their practice following submission of an e-consult request:  I think quite often you need to make a very quick phone call to the patient to clarify a detail or to explain a plan and that is often a problem. They don’t answer the phone, it’s not at a convenient time for them, they’re not expecting it … I wonder whether a more ready way of being able to reply by email. (P4_01/GP) [12] |
|  | **Asynchronous two-way communication** | **Equality:** Increased equity (qualitative) | **CERQual rating:** Low  **CERQual explanation:** Low adequacy  **References:** [13], n=1  **Exemplar data:** e-consultation offered an asynchronous and text-based approach, which was recognised as useful for people who were very anxious, or found face-to-face contact difficult, who had hearing or communication difficulties, and those who struggled to express themselves. [13] |
|  |  | **Safety:** Neutral-increased patient safety (qualitative and quantitative) | **CERQual rating:** Low  **CERQual explanation:** Low adequacy and low coherence  **References:** [1], n=1  **Exemplar data:** Convenience included being able to receive care on their own schedule through asynchronous messaging. There was also evidence that this convenience made patients more likely to seek care when needed. [1] |
|  |  | **Efficiency:** Decreased patient safety (qualitative) | **CERQual rating:** Low  **CERQual explanation:** Low adequacy  **References:** [14], n=1  **Exemplar data:** The extensive text bulk produced by a chat meant the staff lacked an overview. Staff operated the digital communication system for a shift at a time, and the patients did not have a set time limit within which to respond, extending the communication to weeks in some cases. Regardless of whether the nurses followed up their assigned chats or dispatched their ongoing chats after each shift, prolonged chats were considered risky, requiring either multitasking or cases being assessed differently, and thus muddling the response to the patient. [14] |
|  |  | **Efficiency:** Increased workload (qualitative and quantitative) | **CERQual rating:** Low  **CERQual explanation:** Low adequacy  **References:** [15-17], n=3  **Exemplar data:** Some nurses experience that working in the chat is slower than working in the phone service, much due to the asynchronous conversation. They describe that sometimes they can conclude the advice quickly with a patient on the phone. Some nurses had a lot of focus on finalising conversations with patients, and mentioned that it could be stressful to wait for patients’ answers in the chat since patients have 12 h to answer. The conversation became more fragmented in the chat than in the phone and therefore harder to keep the focus on the particular problem and remember all the details about it. [16] |
|  | **Artificial intelligence:** adapting patient questions during query submission | **Efficiency:** Increased workload (qualitative and quantitative) | **CERQual rating:** Low  **CERQual explanation:** Low adequacy  **References:** [7, 17], n=2  **Exemplar data:** Although patients appreciated the potential benefits of an automated history-taking system, they expressed a sense of unease in answering the questions in certain ways, leading to undesired consequences, such as selecting a response alternative that guided them away from the line of questioning that they felt was relevant, with no opportunity to retract. The back-up, that is the possibility in the subsequent chat for correcting any mistakes from the automated history-taking procedure, was considered reassuring but ineffective, duplicating efforts. [17] |
|  | **Artificial intelligence:** Signposting patients to the most appropriate care provider | **Efficiency:** Decreased workload (qualitative and objective) | **CERQual rating:** Low  **CERQual explanation:** Low adequacy, methodological concerns  **References:** [18], n=1  **Exemplar data:** Despite designing the tool with this conservative approach, the most frequent triage disposition was self-care. The majority of these patients did not make further contact with our health system during the subsequent 2 days. This tool may have therefore prevented hundreds of unnecessary encounters. [18] |
|  |  | **Safety:** Decreased patient safety (qualitative) | **CERQual rating:** Low  **CERQual explanation:** Low adequacy, low coherence  **References:** [19], n=1  **Exemplar data:** A total of 40 (21.2%) patients were recommended in their e-consultation to seek urgent/emergency care, but only 16 patients followed this advice. [19] |
|  | **Artificial intelligence:** Prioritising patient queries based on clinical urgency | **Efficiency:** Increased workload (qualitative and objective) | **CERQual rating:** Low  **CERQual explanation:** Low adequacy  **References:** [10], n=1  **Exemplar data:** Staff at a practice using an automated triage algorithm also described the extra work created by ‘overly cautious’ safety mechanisms built into the tool, which meant ‘minor things seem to get flagged up as need-to-be-seen’ (GP1, Pr5, F). [10] |
| **Adopters**  (expected users of OC) | **Security concerns** | **Adoption:** Low adoption by patients (qualitative and quantitative) | **CERQual rating:** Low  **CERQual explanation:** Low adequacy, methodological concerns  **References:** [9, 20], n=2  **Exemplar data:** Public survey respondents were concerned about security (27.5%) and confidentiality (26%) of information that they would need to provide for an online consultation. [20] |
|  | **Visual and cognitive impairments** | **Patient-centeredness:** Decreased patient satisfaction (qualitative) | **CERQual rating:** Low  **CERQual explanation:** Low adequacy  **References:** [21], n=1  **Exemplar data:** The online visual element could be a disadvantage, for example, in a patient with sight problems:  ‘Having a sight problem, I need help to fill this in. It makes things less confidential for me.’ (F, 63 years) [21] |
|  | **Chronic disease** | **Efficiency:** Decreased workload (qualitative and quantitative) | **CERQual rating:** Low  **CERQual explanation:** Low adequacy  **References:** [1, 22, 23] n=3  **Exemplar data:** E-consultation was seen as a simple and secure communication channel with patients, especially with those who have a chronic condition. This enabled a more efficient exchange of information, which was also documented in the electronic patient journal. [23] |
|  |  | **Efficiency:** Decreased costs (qualitative and objective) | **CERQual rating:** Low  **CERQual explanation:** Low adequacy  **References:** [24], n=1  **Exemplar data:** Even greater cost savings are likely to be realized when e-visits are used for the care of patients with chronic diseases such as diabetes and congestive heart failure. The majority of the care a person needs to manage a chronic disease must come directly from the patient. E-visits have the potential to enhance necessary communication between patients who have chronic illnesses and the health care team. [24] |
|  | **Rural areas** | **Adoption:** Low adoption by patients (qualitative and quantitative) | **CERQual rating:** Low  **CERQual explanation:** Low adequacy  **References:** [25, 26], n=2  **Exemplar data:** The utilization of digital primary care in Sweden is highly concentrated in the larger metropolitan areas of Stockholm, Västra Götaland (Gothenburg), and Skåne (Malmö); not shown. In particular, the number of visits by people in Stockholm is around three times higher than in the two other larger cities of Sweden and several times higher than in the rest of the country. [26] |
| **Organisation**  (work needed to implement OC) | **Timely response** | **Efficiency:** Decreased workload (qualitative and objective) | **CERQual rating:** Low  **CERQual explanation:** Low adequacy  **References:** [27], n=1  **Exemplar data:** Doctors said that quick responses lower the amount of double seeking (patients seeking care through multiple channels) [27]. |
|  | **Unclear communication** | **Patient-centeredness:** Decreased patient satisfaction (qualitative) | **CERQual rating:** Low  **CERQual explanation:** Low adequacy, low coherence, methodological concerns  **References:** [9], n=1  **Exemplar data:** No clear documents or communications about how the GP practice is working and how to contact the practice.  Information only available digitally  Number of apps and platforms confusing – not always clear which system GP is using  Language used not easy to understand e.g. “online triage” [9] |
| **Wider system**  (policy context) | Health insurance and payments for services | **Adoption**: High adoption (qualitative and quantitative) | **CERQual rating:** Low  **CERQual explanation:** Low adequacy  **References**: [6], n=1  **Exemplar data:** Patients whose insurers would cover the cost of e-Visits were more likely to use this service. [6] |
|  |  | **Efficiency:** Decreased costs (qualitative and quantitative) | **CERQual rating:** Low  **CERQual explanation:** Low adequacy, methodological concerns  **References:** [22], n=1  **Exemplar data**: OC costs were reimbursed for patients who were insured, in contrast to telephone consultations which were not “billable”. The overall cost of online visits was lower than for office visits. [22] |
