## Appendix 9 for "Understanding how the design and implementation of Online Consultations influence primary care outcomes: Systematic review of evidence with recommendations for designers, providers, and researchers"

**Appendix 9. Outcomes of Online Consultations in primary care (Table 2 – with exemplar data)**

| **Theme** | **Subtheme** |
| --- | --- |
| **Safety** (harm to patients) | **Decreased patient safety (qualitative)**  **Description:** Patient and staff perceptions that OCs worsened patient safety.  **CERQual rating:** High  **References:** [1-17], n=17  **Exemplar data:**  All groups felt that communicating via text led to some loss of communication nuance. While facial expressions and body language were already absent in telephone consultations, cues like tonality were further removed when moving to text-based communication. Staff felt that these cues, in certain situations, provided important ‘between the lines’ context for interpretation of the reported symptoms. [2] |
|  | **Neutral-increased patient safety (qualitative and quantitative)**  **Description:** No quantitative evidence of negative impacts on patient safety, with clinician and patient perceptions that OCs improved patient safety.  **CERQual rating:** High  **References:** [2, 3, 5-8, 18-36], n=25  **Exemplar data:**  There was no difference in apparent treatment failures between the three (telephone, F2F, eVisits) initial encounter types, as measured by antibiotic retreatment with an extension of the initially prescribed antibiotic, or an antibiotic change in therapy within the subsequent 30 days between initial encounter types. There were no statistically different clinical outcomes between the three initial encounter types as evaluated by diagnosis of pyelonephritis within the 30-day follow-up period... There were no hospitalizations or insistences of sepsis within 30 days of initial encounter in any patients. [18] |
| **Timeliness**  (reducing waits and delays) | **Increased access (qualitative and quantitative)**  **Description:** Easier and more convenient for patients to contact their primary care provider, and quicker to communicate with a health professional.  **CERQual rating:** High  **References:** [1-5, 7-9, 11-14, 16, 21-24, 27-29, 31-33, 37-42], n=29  **Exemplar data:**  “We were having a hard time getting through the receptionist at our doctor’s office to make appointments, so [our provider] suggested [virtual visits] as a way to get through to her for things that don’t require us to come into the office” – INTERVIEW P001, F… Virtual visits appeared to improve access to primary care through timely appointments with no booking required… Several survey and interview respondents commented on the difficulty they experienced when attempting to book in-person appointments, such as waiting on the phone with reception or the limited availability of same or next day appointments. Through the virtual visit platform, patients were able to have appointments with their PCP without needing to book them, thereby circumnavigating administrative staff. [23] |
| **Efficiency** (avoiding waste) | **Decreased workload (qualitative and quantitative)**  **Description:** Less work for staff and patients to provide and receive care, respectively.  **CERQual rating:** High  **References**: [1-3, 5-11, 16, 17, 19-27, 29-33, 36, 38-43], n=33  **Exemplar data:**  Advantages for GP offices included reduced phone load, increased efficiency, released time for medical assessments, less crowded waiting rooms and more precise communication. [21] |
|  | **Increased workload (qualitative and quantitative)**  **Description:** More work for staff and patients to provide and receive care, respectively.  **CERQual rating:** High  **References:** [1-3, 5, 6, 8, 9, 11, 13, 15, 21, 22, 24, 27, 29-31, 33, 38-42, 44-49], n=29  **Exemplar data:**  There was a perception that e-consultations could be changing the consultation threshold with patients more likely to complete an e-consultation for issues that may not have been raised through the usual appointment system. This led to a change and possible increase in the workload. This was seen as a different method of access, which brought with it a different type of enquiry: ‘It’s very easy to access and for some patients they may not have brought that particular niggle at all because actually they sort of thought it, we’re not sure, we need this, but because there is this way of e-consulting, then it is another way of coming in to consult with us.’ (CN07) [9] |
|  | **Decreased costs (qualitative and quantitative)**  **Description:** Lower costs for the healthcare system and patients to provide and receive care, respectively.  **CERQual rating:** High  **References:** [1, 3, 6, 7, 9-11, 19, 21, 23, 24, 32, 33, 35, 36, 38, 40, 43, 50, 51], n=20  **Exemplar data:**  After the intervention, the patients with a virtual visit had a lower trend than their matched controls (Can –$3.79, P=.01). The result was an apparent lower expenditure among the virtual visit group at the end of the follow-up period, but again this period was limited. The trends were the same when the outcome was primary care visits rather than costs… Nearly half (48.4%, 193/399) of patients indicated they would have gone to a walk-in clinic if the virtual visit had not been available, 20.3% (81/399) would have had an in-person visit with their doctor or regular place of care, and 10.8% (43/399) would have gone to the emergency department. [24] |
|  | **Increased costs (qualitative and quantitative)**  **Description:** Higher costs for the healthcare system.  **CERQual rating:** High  **References:** [3, 6, 7, 9, 11, 21, 42, 45, 49], n=9  **Exemplar data:**  The average cost for the initial practice response to an e-consultation was £36.28. In context, the national estimates of cost for a standard GP face-to-face consultation is £33.00. The cost was driven mainly by the time needed for a GP to triage the e-consultations and the relatively high proportion of e-consultations that resulted in a face-to-face or telephone consultation with a GP. [49] |
| **Equitable** (variation because of personal characteristics) | **Decreased equity (qualitative and quantitative)**  **Description:** OC use variation based on patient characteristics.  **CERQual rating**: High  **References:** [1, 3, 7-9, 11-14, 16-24, 32-34, 36, 38, 39, 41-45, 47-49, 51-58], n=40  **Exemplar data:**  The experience described through this study is consistent with other international work showing that telehealth risks increasing inequity. [7] |
|  | **Increased equity (qualitative)**  **Description:** OCs helped patients communicate with their primary care providers who had previously struggled due to their personal characteristics.  **CERQual rating**: High  **References:** [1, 3, 7, 9, 11, 12, 14, 16, 21-23, 29, 38, 39, 41, 42, 45, 53, 57], n=19  **Exemplar data:**  Patients and GP staff commented on the benefits of providing individuals with different means of communicating with their GP. This flexibility was felt to ensure that patients felt they had a means of communicating in a way that suited them and they were comfortable with. [22] |
| **Patient-centredness** (care that is respectful and responsive) | **Decreased patient satisfaction (qualitative)**  **Description:** Negative patient experiences of using OCs.  **CERQual rating:** High  **References:** [1, 3, 11-13, 16, 22, 23, 26, 29, 33, 38, 41], n=13  **Exemplar data:** Some patients expressed a dislike of the asynchronous interaction offered by the platform: ‘If you’re under stress because you’re poorly, I’d prefer to speak to a human!’ (F, 47 years) [16] |
|  | **Increased patient satisfaction (qualitative and quantitative)**  **Description:** Positive patient experiences of using OCs.  **CERQual rating:** High  **References:** [1, 3, 4, 6-9, 11-14, 16, 17, 20-24, 26, 29, 30, 33, 35-42, 50], n=31  **Exemplar data:**  81% of the patients would recommend this DPHC concept to others (Q18), and a majority of the patients (72%) experienced equal or greater satisfaction with the service compared with a physical visit (Q16). Among those patients who were more satisfied with the DPHC concept (n = 36; 26%), their main reason was availability (Q17, Table 1). These responses correspond well with the overall satisfaction with DPHC (Md, 8.0; IQR, 6-9; Q19) [4] |

13.	Health Innovation Manchester. GM Digital First Primary Care: Patient and public insights: Workshop results. Report. Manchester: Health Innovation Manchester, 2021 Sept 2021. Report No.: 1 Contract No.: 1 Sept.
