## Appendix 10 for "Understanding how the design and implementation of Online Consultations influence primary care outcomes: Systematic review of evidence with recommendations for designers, providers, and researchers"

**Appendix 10. How outcomes of Online Consultations in primary care are influenced by system design and implementation (Table 3 – with exemplar data)**

| **Theme** | **OC design feature or implementation** | **Outcome (from Table 2)** | **CERQual rating, references, and exemplar data** |
| --- | --- | --- | --- |
| **Condition** (illness OC is used for) | **Decreased complexity of query**  **Description:** Patient queries are straightforward and easy to resolve e.g. administrative tasks, minor acute illnesses, and prescription requests. | **Efficiency:** Decreased workload (qualitative and quantitative)  **Efficiency:** Decreased health costs (qualitative and quantitative) | **CERQual rating:** High  **References:** [1-11], n=11  **Exemplar data:**  The digital communication system also procured efficiency such as settling simple cases easily, relieving the telephone service of those minor issues that can be resolved by a single message turnaround… The findings suggested that the digital communication service was best suited to less complex matters… [3] |
|  | **Increased complexity of query**  **Description:** Patient queries are not straightforward and easy to resolve e.g. multiple ill-defined symptoms. | **Efficiency:** Increased workload (qualitative)  **Efficiency:** Increased health costs (qualitative and quantitative) | **CERQual rating:** High  **References:** [1, 3-5, 8, 12, 13], n=7  **Exemplar data:**  For patients presenting with a complex or new set of symptoms clinicians usually felt the need to talk to the patient directly: ‘When someone says, “I have felt unwell for 3 weeks with headache, dizziness, limb aches, vision’s blurred” it’s just impossible to actually disentangle that with an e-consult, you’ve got to see them.’ (CN17) [4] |
| **Technology** (material properties of OC) | **Multiple choice questionnaires (MCQ)**  **Description:** Patients describe their query by completing questionnaires and selecting their answers from a list. | **Efficiency:** Increased workload (qualitative) | **CERQual rating:** High  **References:** [3-5, 8, 11, 14-21], n=13  **Exemplar data:**  Some of the nurses explained that they needed to ask more questions in the chat system than in a phone call, because they did not trust the information to the same degree in the chat system as on the phone. One of the nurses explains:  “It may not belong [in the patient’s case], while in the chat they only get the standard questions in the questionnaire and then they have answered yes to many different things, which they probably have [symptoms of] but may not at the moment or do not relate to what they are looking for help right now. So then I have to dig more into it”. [18] |
|  |  | **Patient-centredness:** Decreased patient satisfaction (qualitative) | **CERQual rating:** High  **References**: [3, 5, 11, 15-17, 19-21], n=9  **Exemplar data:**  Although patients appreciated the potential benefits of an automated history-taking system, they expressed a sense of unease in answering the questions in certain ways, leading to undesired consequences, such as selecting a response alternative that guided them away from the line of questioning that they felt was relevant, with no opportunity to retract. The back-up, that is the possibility in the subsequent chat for correcting any mistakes from the automated history-taking procedure, was considered reassuring but ineffective, duplicating efforts. [19] |
|  | **Free-text input**  **Description:** Patients describe their query using unstructured text. | **Efficiency:** Decreased workload (qualitative and quantitative)  **Safety:** Increased patient safety (qualitative) | **CERQual rating:** High  **References:** [1, 14, 16, 22-25], n=7  **Exemplar data:**  A physician at Group Health sends his patients a secure message several days before their appointment asking for their concerns. This improves the efficiency of office visits: “half the time they’ve written the history of present illness. I just copy and paste it into the EMR.” [23] |
|  | **Asynchronous two-way written communication**  **Description:** Patients and staff are able to send written messages to each other at different times. | **Efficiency:** Decreased workload (qualitative and quantitative) | **CERQual rating:** High  **References:** [2, 14, 22, 25-27] n=6  **Exemplar data:**  The platform was perceived to provide a unique value through the asynchronous chat, as clinical decisions could be communicated with several short messages without excessive conversation. [14] |
|  | **Non-integration with core software systems**  **Description:** OC systems that operate separately from other software used by the primary care provider. | **Efficiency:** Increased workload (qualitative) | **CERQual rating:** High  **References:** [3-6, 8, 13-16, 18, 24, 28, 29], n=13  **Exemplar data:**  The processing of the e-consult is usually more time consuming for administrative staff and GPs than an email. That e-consults could not be directly linked to patients’ notes [EHR] without administrative staff input was perceived to be a major limitation of e-consults. [16] |
| **Adopters** (expected users of OC) | **Female gender**  **Description:** Female patients. | **High adoption** (qualitative and quantitative)  **Equitable:** Decreased equity (perceived and objective) | **CERQual rating:** High  **References:** [5, 6, 8, 10, 12, 15, 16, 25-28, 30-44], n=26  **Exemplar data:**  Female patients accounted for 88% of eVisits. [10] |
|  | **Lower age**  **Description:** Younger patients. | **High adoption** (qualitative and quantitative)  **Equitable:** Decreased equity (qualitative and quantitative) | **CERQual rating:** High  **References:** [5-8, 10-13, 16, 21, 27, 28, 30, 31, 36-42, 44-48], n=26  **Exemplar data:**  On average, eVisit adopters are younger… On average, a patient's odds of eVisit use is 2.6% lower when the patient is 1 year older. [30] |
|  | **Native speakers**  **Description:** Patients who are native speakers of the official language in the country they live. | **High adoption** (qualitative and quantitative)  **Equitable:** Decreased equity (qualitative and quantitative) | **CERQual rating:** High  **References:** [5, 8, 20, 26, 42, 46, 49], n=7  **Exemplar data:**  Those who do not have English as a first language or speak English are unable to use the tools – rely on family members or community to support with translation. [20] |
|  | **High socioeconomic status**  **Description:** Patients with higher levels of income and education. | **High adoption** (qualitative and quantitative)  **Equitable:** Decreased equity (qualitative and quantitative) | **CERQual rating:** High  **References:** [5-8, 20, 26, 36, 40, 41, 50, 51], n=11  **Exemplar data:**  912 participants who had completed a virtual visit provided responses to the survey. Participants were primarily Caucasian (78.5 %), married (71.2 %), highly educated (79.8 % had a post-secondary degree), and of high family income (55.8 % had a family income of over $90,000). [26] |
|  | **Mental health conditions**  **Description:** Patients with a mental health diagnosis. | **Timeliness:** Increased access (qualitative)  **Equitable:** Increased equity (qualitative)  **Patient-centredness:** Increased patient satisfaction (qualitative and quantitative) | **CERQual rating:** High  **References:** [5, 6, 11, 13, 15, 19, 21, 26], n=8  **Exemplar data:**  Some [staff] also mentioned particular conditions and situations for which webGP may be advantageous, such as problems with mental health: ‘There’s one really good example … It was a girl with anxiety … She got embarrassed easily and stumbled over her words and that was her barrier to actually coming and discussing it in the first place.’ (P2_02/GP) [15] |
|  | **Verbal communication difficulties**  **Description:** Patients with difficulty communicating verbally e.g. those with hearing loss. | **Timeliness:** Increased access (qualitative)  **Equitable:** Increased equity (qualitative)  **Patient-centredness:** Increased patient satisfaction (qualitative and quantitative) | **CERQual rating:** High  **References:**  [1, 4, 5, 11, 13, 40, 51], n=7  **Exemplar data:**  There were certain health conditions where a non-verbal form of communication was easier:  ‘For a deaf person it is marvellous to be able to communicate without using a voice phone.’  (Male [M], 83 years) [40] |
|  | **Physical barriers to attending in-person appointments**  **Description:**  Patients cannot easily attend in-person appointments e.g. due to physical disabilities, living far from their primary care provider, work commitments, or care responsibilities. | **Timeliness:** Increased access (qualitative)  **Equitable:** Increased equity (qualitative)  **Patient-centredness:** Increased patient satisfaction (qualitative and quantitative) | **CERQual rating:** High  **References:** [5-8, 11, 15, 46, 47], n=8  **Exemplar data:**  Others favoured the online format and remote consultation style; they used the system as it was difficult to visit the practice due to disabilities, illness or working commitments… [8] |
|  | **Preference for traditional consulting methods**  **Description:** Staff and patients believe in-person consultations are the gold standard. | **Low adoption** (qualitative) | **CERQual rating:** High  **References:** [5, 7, 13, 23, 46, 50-52], n=8  **Exemplar data:**  Barriers included difficulties in making patients aware of the option to use an alternative to the face-to-face consultation and subsequently getting them to engage with these alternatives when the face-to-face consultation was still seen as the ‘gold standard’. [13] |
| **Organisation** (work needed to implement OC) | **Lack of OC promotion**  **Description:** Patients are not effectively informed that OCs are available for them to contact their primary care provider. | **Low adoption** (qualitative and quantitative) | **CERQual rating:** Moderate  **References:** [1, 5, 25, 50, 51], n=5  **Exemplar data:**  Concern that many people are unaware of the digital technologies and apps (AskmyGP, NHS), highlighting the need to raise awareness and provide support as people do not always have technologies or access to family or community groups to support them (Shielding group, BAME group). [51] |
|  | **Timely response**  **Description:** Primary care providers respond quickly to patients’ OC queries. | **Patient-centredness:** Increased patient satisfaction (qualitative and quantitative)  **Timeliness:** Increased access (qualitative) | **CERQual rating:** High  **References:** [8, 15, 16, 20, 26, 28, 53], n=7  **Exemplar data:**  Other users [patients] echoed a favour for the speed of response: “When I message them today they will call me straight away” “I like how every time I send a request I get an answer straight away” [53] |
|  | **Non-integration with daily workflows**  **Description:** Primary care provider does not coherently plan OCs into their work processes e.g. by not scheduling clinician time to deal with OCs, or not diverting as much incoming patient demand via OCs as possible. | **Efficiency:** Increased workload (qualitative and quantitative) | **CERQual rating:** High  **References:** [3-5, 7, 13-15, 17, 21, 23, 28, 29, 31], n=13  **Exemplar data:**  …providers lamented that electronic communication made the workday longer. As the number of electronic communications with patients increased, several groups tried to cut down on the number of office visits but, in most cases, the number of office visits did not decrease very much. Electronic communication therefore was often added work to a full day of office visits. [23] |
|  | **Sufficient resources allocated to implementing OCs**  **Description:** Adequate training, staff, and facilities are available to conduct OCs. | **Efficiency:** Decreased workload (qualitative) | **CERQual rating:** High  **References:** [3, 6, 7, 14, 17, 21, 23, 28], n=8  **Exemplar data:**  “We need to give them [the right training] so that they feel more confident to say, 'Okay, I diagnose you,' for example, 'You have tonsillitis. I felt confident making sure that you had to open your mouth properly and I could see your tonsils, even though it was through a phone, and therefore I could diagnose you effectively as I would have done in a face-to-face consultation.' So, people overcompensate that time and therefore they spend a little bit longer, and therefore the efficiency gained that might have been understood through the virtual consultation might not be realised.” Stakeholder [6] |
|  | **Lack of continuity**  **Description:** OC query is not dealt with by a known or preferred physician | **Patient-centredness:** Decreased patient satisfaction (qualitative) | **CERQual rating:** Moderate  **References:** [6, 11, 28, 35, 53], n=5  **Exemplar data:** When online consultations could potentially be reviewed and answered by any GP, both staff and patients noted further unintended consequences negatively impacting continuity of care: ‘[Online consultations are] very much a move from […] a nice doctor–patient relationship [...] we try and maintain continuity, but that’s difficult with this system […] often other people will pick up calls that are meant for you or the patients don’t specifically ask for you.’ (GP1, Pr1, F) [11] |
| **Wider system** (policy context) | **Government policy**  **Description:** Policies mandating OC usage e.g. by increasing digital modes of contact with primary care in general or minimising in-person contact during the COVID-19 pandemic. | **High adoption** (qualitative and quantitative) | **CERQual rating:** High  **References:** [6, 29, 36, 46, 54, 55], n=6  **Exemplar data:**  [The] significant increase of e-visits in Denmark is the result of a policy to increase the supply of e-visit by mandating GPs to offer e-visits. Whether the use of the online channel is optional for the patients, all GPs must offer the service. [36] |
|  | **Lack of financial support**  **Description:** No external funding available to pay ongoing costs of OCs. | **Low adoption** (qualitative and quantitative) | **CERQual rating:** Moderate  **References:** [3, 5, 7, 8, 46], n=5  **Exemplar data:**  None of the 36 practices took up the system after the pilot, which would have involved paying market prices for the software. However, 13 practices were interested in continuing to use the system if costs were paid for by alternative funding sources. [8] |

20.	Health Innovation Manchester. GM Digital First Primary Care: Patient and public insights: Workshop results. Report. Manchester: Health Innovation Manchester, 2021 Sept 2021. Report No.: 1 Contract No.: 1 Sept.
